# RANKL inhibition spatially rewires the microenvironment of luminal breast cancer

**DOI:** 10.64898/2026.07.08.26355790

**Authors:** Mario Rodriguez-del Collado, Andrea Vethencourt, Alexandra Barranco, Jaime Martinez de Villarreal, David Valcárcel-Linares, Eva Maria Trinidad, Eduard Dorca, Gonzalo Soria-Alcaide, Maria Jimenez, Eduardo J. Caleiras, Maria Gomez, Carlos Garrido, Orlando Dominguez, Gema Perez-Chacon, Marina Ciscar, Elvira Purqueras, Gonzalo Gomez, Elena Piñeiro-Yañez, Ander Urruticoechea, Isaac Subirana, Javad Noorbakhsh, Jeffrey H. Chuang, Anna Petit, Maria Teresa Soler-Monso, Anna Gumà, Sonia Pernas, Catalina Falo, Eva Gonzalez-Suarez

## Abstract

Hormone receptor-positive, HER2-negative breast cancers are often poorly infiltrated by immune cells and derive limited benefit from current immunotherapy strategies. Here, using paired tumour samples from the randomised window-of-opportunity D-BIOMARK trial (NCT03691311), we investigated the immunomodulatory effects of denosumab in early luminal breast cancer. Short-term preoperative denosumab reduced tumour-cell proliferative transcriptional programs and immunosuppressive features of the local tumour microenvironment, enhancing innate and adaptive immune activation and altering circulating cytokine profiles. High-resolution spatial transcriptomics revealed coordinated remodelling of tumour, immune, fibroblast and endothelial compartments after treatment. Denosumab reduced immune–tumour spatial separation and enhanced T cell activation, accompanied by a shift from matrix-associated tumour programs towards increased tumour–T cell communication. Copy number-informed tumour-state inference further identified a reduced representation of genomically complex, immune-poor tumour subclones after treatment. Together, these findings identify denosumab as a modulator of tumour–microenvironment crosstalk and support RANKL blockade as a strategy to render immune-poor luminal breast tumours more permissive to immune engagement.

## INTRODUCTION

Immunotherapy has reshaped the treatment of triple-negative breast cancer, but its benefit in hormone receptor-positive, human epidermal growth factor receptor 2 (HER2)-negative disease remains limited. This is a major clinical challenge, as luminal tumours represent the most common form of breast cancer (Vonderheide et al., 2017). Beyond reduced lymphocytic infiltration, these tumours display immunosuppressive features that include stromal remodelling, macrophage-enriched niches, and fibroblast-mediated immune exclusion (Goldberg et al., 2021). Together, these properties contribute to limited sensitivity to immune checkpoint blockade. However, immune exclusion is not only determined by cell abundance, but also by the spatial organisation of immune, stromal and tumour compartments, which remains incompletely understood in luminal tumours.

The receptor activator of nuclear factor-κB (RANK) and its ligand RANKL constitute a pharmacologically targetable pathway, with the anti-RANKL antibody denosumab approved for osteoporosis and for the prevention of skeletal-related events in patients with bone metastases (Pérez-Chacón et al., 2025; U.S. Food and Drug Administration, 2025). In the mammary epithelium, RANK signalling contributes to hormone-driven proliferation, survival and tumour initiation, while in established breast cancer it has been implicated in recurrence, metastasis and immune suppression (Ciscar et al., 2023; Gómez-Aleza et al., 2020; Pérez-Chacón et al., 2025; Yoldi et al., 2016), reinforcing the therapeutic potential of RANKL blockade in breast cancer. Consistent with this, preclinical experiments and D-BEYOND clinical data from the D-BEYOND study showed that genetic or pharmacological inhibition of RANK signalling increased stromal tumour-infiltrating lymphocytes (TILs) and reinforced anti-tumour immune responses in luminal tumours (Gómez-Aleza et al., 2020). Similarly, the randomised D-BIOMARK window-of-opportunity trial, which included an untreated control arm, showed that a short course of denosumab increased stromal TILs in early HER2-negative breast cancer (Vethencourt et al., 2025). These findings support an immune-enhancing role for denosumab in breast cancer. However, the transcriptional, cellular and spatial mechanisms underlying this effect remain unresolved, particularly in luminal tumours, where modest changes in immune composition may be missed or misinterpreted when assessed by histology alone (Zhu et al., 2019).

Here, using paired biopsy and surgical tumour samples from the randomised D-BIOMARK trial, we define the transcriptomic and immune-compositional effects of preoperative denosumab in early luminal breast cancer. We then use Visium HD spatial transcriptomics, proximity analyses, cell–cell communication inference and copy-number-informed tumour-state analysis to determine how these changes are organised across the tumour microenvironment (TME). This enables the identification of coordinated multicellular responses, spatial immune niches and tumour-stroma interactions associated with RANKL blockade. Our findings identify denosumab as a modulator of the TME interactions in luminal breast cancer and provide a framework for understanding how RANKL blockade may render poorly infiltrated tumours more permissive to immune engagement.

## RESULTS

### Preoperative denosumab transcriptionally reshapes the TME in early luminal breast cancer

To assess the biological impact of preoperative denosumab in luminal breast cancer, we analysed paired diagnostic biopsy and surgical luminal tumour specimens from the D-BIOMARK trial, in which patients were randomised 2:1 to receive two preoperative doses of denosumab (120 mg subcutaneously, 7 days apart; Fig. 1A). In luminal tumours, the percentage of stromal TILs in tissue samples increased after denosumab treatment (n=31, p = 0.005; Fig. 1B), consistent with previous findings from D-BEYOND. However, TILs also tended to increase in the control arm (n= 17, p = 0.18; inter-p = 0.72; Fig. 1B; Supplementary Table S1), indicating that TILs accumulation alone does not fully capture denosumab-associated effects.

**Figure 1.**
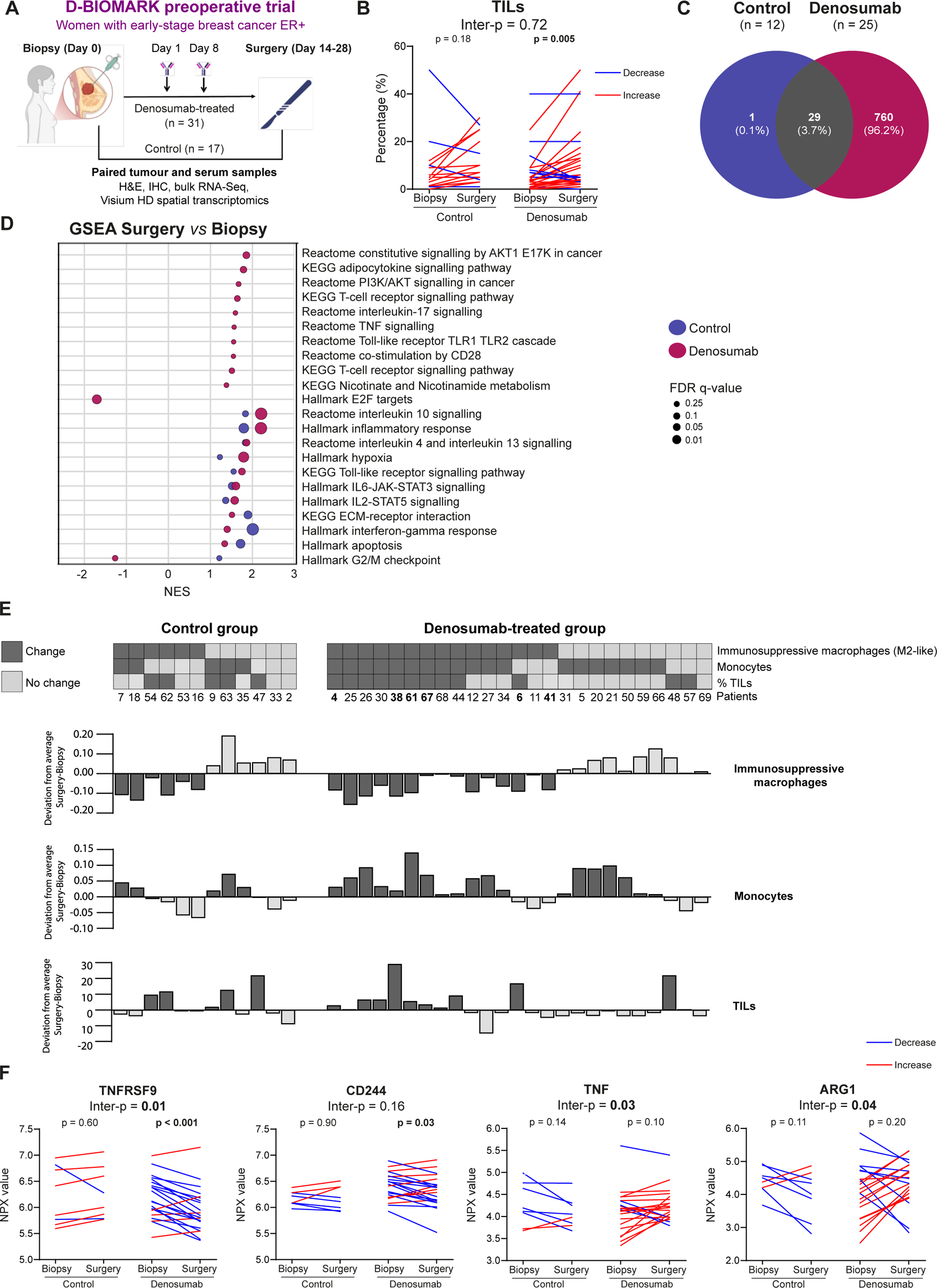
Preoperative denosumab shows coordinated immunomodulatory effects in the tumour microenvironment and in circulation in early luminal breast cancer. **A.** Study design of the D-BIOMARK window-of-opportunity clinical trial (NCT03691311). Women with early-stage ER-positive, HER2-negative breast cancer were randomized 2:1 to receive two preoperative subcutaneous doses of denosumab (120 mg, 7 days apart; n = 31) or control (n = 17). Paired tumour tissue and serum samples were collected at baseline and surgery and analysed using complementary profiling approaches. **B.** Percentage of stromal tumour-infiltrating lymphocytes (TILs) in paired biopsy and surgery samples from control and denosumab-treated patients with early luminal breast cancer. **C.** Overlap of differentially expressed genes (DEGs) identified by paired bulk RNA-Seq analysis in control (n = 12) and denosumab-treated (n = 25) tumours. DEGs were defined by adjusted p < 0.05. **D.** Gene set enrichment analysis (GSEA) comparing surgery *versus* biopsy samples in control and denosumab-treated tumours. Selected pathways are shown; dot colour indicates treatment arm, and dot size indicates FDR q-value. NES positive values indicate enrichment at surgery and negative values indicate enrichment at biopsy. **E.** Patient-level classification of denosumab-associated immune remodelling based on coordinated changes in immunosuppressive macrophages, monocytes and TILs. The heat map indicates whether each patient met the prespecified criteria for change (decrease in immunosuppressive macrophages measured by CIBERSORT, increase in monocytes measured by CIBERSORT, and increase in TILs measured by H&E); bar plots show each patient’s deviation from the mean surgery-biopsy change in the control arm. **F.** Levels of TNFRSF9, CD244, TNF and ARG1 immune-related proteins in paired serum samples from patients in the control (n = 8) and denosumab-treated (n = 21) groups of the D-BIOMARK clinical trial. See Extended Data Figure 1-3 and Supplementary Tables S1-S2

Analyses of immune composition by immunohistochemistry (IHC) revealed an increase of cytotoxic T cells (p = 0.01) and CD68+ macrophages (p = 0.04) in denosumab-treated tumours, whereas control tumours showed increased regulatory T cells (p = 0.01), helper T cells (p = 0.006) and B lymphocytes (p = 0.04). MUM1+ plasma cells, PD1+ cells and PD-L1 expression remained unchanged. Among the immune populations assessed, regulatory T cells showed the clearest divergence between treatment arms, although this did not reach significance in the between-arm comparison (inter-p = 0.08) (Extended Data Fig. 1A; Supplementary Table S1).

To define these changes at the transcriptional level, we performed bulk RNA sequencing (RNA-Seq) on paired tumours from 37 patients (experimental arm, n = 25; control arm, n = 12). The number of differentially expressed genes (DEGs) between biopsy and surgery was 26-fold higher in the denosumab arm than in the control arm (789 versus 30 DEGs, respectively, adjusted p < 0.05; Fig. 1C; Supplementary Table S2), indicating a broad molecular response to denosumab. Modulated genes included cytokines and chemokines, such as *CCL3, CCL4*, *IL1B*; early oestrogen-response genes, including *FOS, CCN1*, *JUNB;* and metabolic and mitochondrial genes, including *ADIPOQ* and *MT-TC.* Gene set enrichment analysis (GSEA) showed that pathways selectively enriched in the experimental arm included “TNF signalling”, “T cell receptor signalling”, “B cell receptor signalling” and “NOD-like receptor signalling”. By contrast, proliferative programmes, such as “E2F targets” and “DNA replication”, were reduced in the experimental arm (Fig. 1D; Supplementary Table S2). Notably, “G2/M checkpoint” was downregulated in the experimental arm but increased in the control arm (Fig. 1D, Supplementary Table S2).

The activity of some pathways increased in both groups, but larger changes were observed in denosumab-treated tumours, including “Interleukin-10 signalling”, “Inflammatory response” and “Toll-like receptor signalling”. By contrast, other pathways such “Extracellular matrix receptor interaction” showed relatively greater changes in controls (Fig. 1D; Supplementary Table S2). To resolve these patterns at the individual-patient level, we applied single-sample GSEA (ssGSEA). Most significant changes were restricted to the experimental arm, including upregulation of immune-related programmes, such as “Inflammatory response”, “Interferon-γ response”, “TNF signalling” and “Toll-like receptor signalling”, together with downregulation of metabolic pathways, including “Respiratory electron transport” and “Mitochondrial biogenesis” (Extended Data Fig. 1B; Supplementary Table S2). The pathways “Nicotinate and nicotinamide metabolism” (inter-p = 0.01) and “Interleukin-17 signalling” (inter-p = 0.07) involved in T cell functionality showed the most divergent behaviour between arms (Extended Data Fig. 1B; Supplementary Table S2). These analyses further support the immunomodulatory effects driven by denosumab, extending beyond differences in immune abundance between biopsy and surgical specimens.

### Denosumab modulates myeloid and lymphoid populations in the TME

To gain insight into denosumab-associated changes in the composition of the immune microenvironment, we performed immune deconvolution using CIBERSORT (Newman et al., 2015). Consistent with the increase in TILs assessed by H&E and IHC, denosumab-treated tumours showed increased lymphocytes (here referring to the combined lymphoid and NK-cell populations inferred by CIBERSORT as TILs; p = 0.03), T cells (p = 0.03) and CD4+ T cells (p = 0.01), whereas no changes were observed in untreated tumours (Extended Data Fig. 1C; Supplementary Table S1). Regulatory T cells decreased in the experimental arm (p = 0.03) but not in controls (p = 0.18), consistent with findings in the D-BEYOND trial (Gómez-Aleza et al., 2020). No significant changes in CD8+ T cells were detected by CIBERSORT in either arm.

Denosumab was also associated with changes in the myeloid compartment. Immunosuppressive macrophage-related scores decreased after treatment (p = 0.02), whereas monocyte-related scores increased (p = 0.001). Total macrophages (p = 0.004) and neutrophils (p = 0.02) also decreased in the experimental arm, but not in controls. Between arm comparisons further supported significant differences for monocytes (inter-p = 0.04), total macrophages (inter-p = 0.04) and plasma cells (inter-p = 0.02) (Fig. 1D; Supplementary Table S1). Notably, nine denosumab-treated tumours showed a concurrent increase in TILs assessed by H&E, and monocyte-associated signatures together with a reduction in immunosuppressive macrophages, both inferred by CIBERSORT, whereas none of the untreated tumours exhibited this coordinated immune remodelling (Fig. 1E). Together, these findings indicate that preoperative denosumab is associated with reduced macrophage-associated immunosuppressive features and enhanced T cell infiltration in early luminal breast cancer.

Given the biological and clinical relevance of molecular subtype and menopausal status in hormone receptor-positive breast cancer, subgroup analyses were performed. These immune-related changes were more pronounced in premenopausal patients, in whom increased lymphocytes- and monocyte-related signatures were accompanied by reductions in immunosuppressive macrophages, total macrophages and regulatory T cells. Significant between arm differences were observed for CD4+ T cells, plasma cells, regulatory T cells, immunosuppressive macrophages, total macrophages and monocytes (Extended Data Fig. 2). Likewise, luminal B-like tumours showed increases in lymphocyte signatures and monocytes, together with reductions in immunosuppressive and total macrophages, whereas luminal A-like tumours showed fewer changes overall, except for increased monocyte signatures and reduced neutrophils (Extended Data Fig. 3).

Individual-patient analyses further revealed that coordinated changes in immune populations were selectively enriched following denosumab treatment. These findings support a model in which denosumab remodels the TME, promoting concerted reorganization across lymphoid and myeloid compartments, particularly in tumours from premenopausal patients and the luminal B-like subtype.

### Preoperative denosumab enhances systemic levels of pro-inflammatory cytokines in patients with luminal breast cancer

Given the general pro-inflammatory environment imposed by RANK pathway inhibition, we next assessed whether the effects observed in the TME were translated into systemic changes. Proteomic profiling of immune-oncology related proteins in serum samples collected at biopsy and surgery from 29 patients revealed a marked reduction in circulating levels of the immune modulators TNFRSF9 (also known as CD137 or 4-1BB) and CD244, whereas no changes were observed in the control arm. Moreover, the pro-inflammatory cytokine TNF and the immunoregulatory enzyme ARG1 showed divergent patterns between the denosumab-treated (increase) and control arms (decrease) (Fig. 1F; Supplementary Table S1). These changes indicate that denosumab-associated immune modulation is not restricted to the TME, but it is also accompanied by systemic immune changes.

### Spatial transcriptomics reveals multicompartment remodelling after denosumab

To investigate how the denosumab response is spatially organised within the TME, we performed Visium HD spatial transcriptomics on paired tumour samples from six denosumab-treated patients. We prioritised cases from the luminal B-like subtype that had shown coordinated remodelling across lymphoid and myeloid compartments in the bulk analyses (patient (ptx) 4, ptx 6, ptx 38, ptx 61 and ptx 67), together with ptx 41, who only showed a small reduction in immunosuppressive macrophages (Fig. 1E). To maximise the ability of the spatial analyses to capture breast cancer heterogeneity and the biology associated with the immune changes described above, we selected paired tumour samples representing diverse clinical and pathological features. These included samples from three cases with increases greater than 10% in TILs after treatment and three with increases of less than 10%; three invasive carcinomas of no special type (NST) and three other histological subtypes (comprising one mixed micropapillary/NST, one solid papillary, and one lobular carcinoma); and three premenopausal and three postmenopausal patients (Supplementary Table S3). PAM50 intrinsic subtype classification confirmed a luminal B profile in most diagnostic biopsies, with only one case classified as HER2-enriched. RANK and RANKL expression was generally low across the cohort, although ptx 61, and ptx 67 showed tumour cell RANK positivity (H-score > 10), whereas stromal RANK expression was detected in ptx 4 and ptx 61 (H-score > 10) (Supplementary Table S3).

Visium HD data quality varied across cases, with ptx 4, ptx 41 and ptx 67 showing a higher proportion of spots with lower unique molecular identifier (UMI) counts than ptx 6, ptx 38 and ptx 61 (Supplementary Table S3). To increase molecular signal robustness while reducing technical sparsity, downstream analyses were performed using 16-μm aggregated spots, according to the predefined strategy described in the Methods. Tumour and non-invasive breast regions were first manually annotated by a certified pathologist, whereas stromal and immune identities were then assigned by robust cell type decomposition (RCTD) using ER-positive breast cancer single-cell and normal breast single-nucleus reference atlases (Kumar et al., 2023; Pal et al., 2021)(Fig. 2A; Extended Data Fig. 4A, Methods). This resolved tumour cells, fibroblasts, myeloid cells, endothelial cells and T cells, together with less abundant populations such as B cells, adipocytes, lymphatic cells, mast cells, perivascular cells and plasmablasts. Tumour-mixed and normal breast-mixed categories were defined to capture spots with mixed epithelial–stromal composition but were excluded from downstream inferential analyses. Marker-gene expression supported the final annotations (Extended Data Fig. 4B).

**Figure 2.**
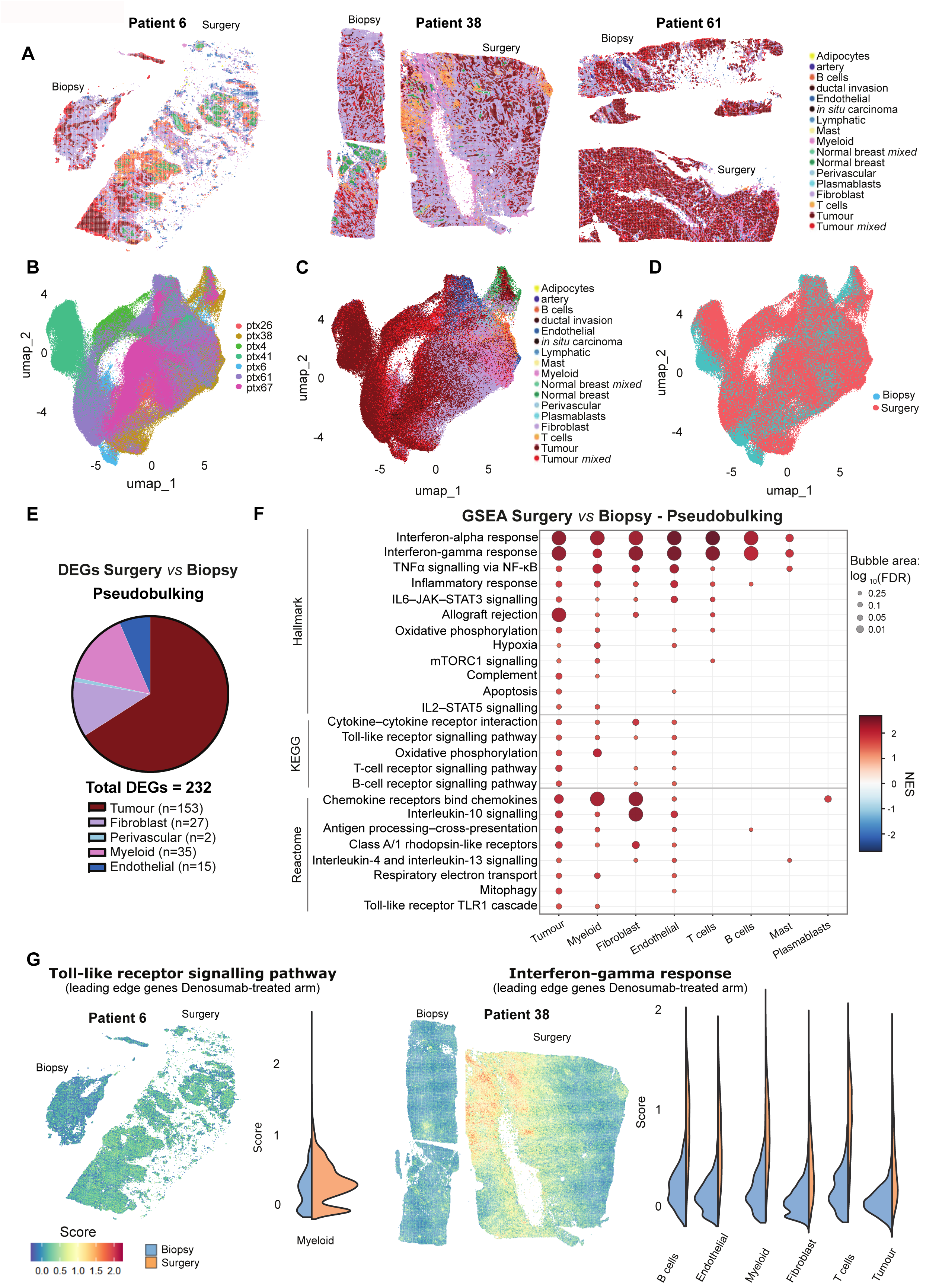
Visium HD resolves multi-compartment transcriptional remodelling after denosumab. **A**. Spatial maps of annotated Visium HD spots in paired biopsy and surgery sections from patients 6, 38 and 61. Cell identities were assigned by integrating pathology-guided annotation of tumour and normal breast regions with RCTD deconvolution of stromal and immune compartments. Tumour-mixed and normal breast-mixed spots denote mixed epithelial-stromal profiles and were excluded from downstream inferential analyses. **B-D**. UMAPs of the integrated Visium HD dataset coloured by patient (**B**), final cell identity (**C**) and time point (**D**). **E**. Distribution of significant pseudobulk DEGs comparing surgery *versus* biopsy samples detected across annotated cell populations (total n = 232); the pie chart reports the contribution of each compartment. **F**. Pseudobulk GSEA comparing surgery *versus* biopsy samples across annotated compartments. Dot colour indicates NES and dot size indicates FDR-adjusted significance. Positive NES values indicate post-treatment enrichment, and negative NES values indicate pretreatment enrichment. Pathways are grouped by Hallmark, Reactome and KEGG collections. **G**. Spatial projection of selected denosumab-associated inflammatory signatures (selecting genes from the leading edge) onto Visium HD sections, with compartment-specific score distributions for the indicated cases and signatures. Colours indicate signature score and violin plots compare biopsy and surgery distributions. See Extended Data Figure 4-6 and Supplementary Tables S3-S6.

Once spot-level annotations were established for all patients, an integrated analysis across the entire cohort was performed, enabling comprehensive transcriptional characterization of all cell types and the identification of the global effects of denosumab treatment across the dataset. Overall, there was good concordance across patients, except for ptx 41, a solid papillary carcinoma lacking stromal TILs, whose tumour population segregated from the remaining tumours (Fig. 2B-C). Pre- and post-treatment samples largely overlapped in the integrated embedding, although substantial inter-patient differences in tissue composition were evident. Samples from ptx6 and ptx38 were enriched for fibroblasts, while ptx41 and ptx67 were dominated by tumour spots and showed sparse immune cell representation, consistent with their low TIL abundance (Fig. 2A–D; Extended Data Fig. 4C; Supplementary Table S3). Despite this heterogeneity, paired pseudobulk analysis identified a post-treatment transcriptional response across multiple tissue compartments, with the strongest signal in tumour cells and similar effects in myeloid, fibroblast and endothelial populations (Fig 2E-F; Supplementary Table S4). The tumour compartment showed the largest number of significant DEGs (n = 153), followed by myeloid (n = 35), fibroblast (n = 27) and endothelial cells (n = 15), whereas perivascular cells showed minimal changes (n = 2) (Fig. 2E).

As an assessment of dataset representativeness, we compared spatial pseudobulk profiles from Visium HD with bulk RNA-Seq data from all denosumab-treated patients. Tumour cells showed the largest absolute overlap with the bulk dataset, with 14 shared differentially expressed genes, consistent with a dominant tumour-intrinsic contribution to the bulk signal. Notably, endothelial cells showed the strongest relative concordance, with 5 of 15 endothelial DEGs overlapping with bulk RNA-Seq, highlighting vascular activation and extracellular matrix remodelling as reproducible non-tumour features of the post-treatment state. Myeloid cells also showed concordance with the bulk response, with 5 of 35 DEGs overlapping, whereas fibroblast overlap was more modest and the perivascular signal remained too limited for strong inference. The observed concordance between platforms supported the ability of the spatial transcriptomic dataset to capture the major transcriptional features of the matched tumour samples, while enabling compartment-level resolution of the post-treatment response (Supplementary Table S4).

GSEA further supported a coordinated post-treatment inflammatory and immune-regulatory response across the tumour microenvironment, also consistent with the bulk transcriptomic changes observed in the whole cohort: with 148 out of the 154 signatures identified in the spatial being present in the experimental arm (Fig. 2F; Supplementary Table S4). Across multiple compartments, enriched programmes included “Interferon gamma response”, “TNFα signalling via NF-κB” and “Cytokine–cytokine receptor interaction”. In tumour cells, these changes extended to “Antigen processing cross presentation” and “Allograft rejection” programmes, and were supported by induction of *FOS, EGR1* and *DUSP1*, consistent with activation of immediate-early signalling networks. Myeloid cells showed a related response, with enrichment of “Interferon alpha”, “Interferon gamma response”, “Inflammatory response” and “Hypoxia”, supported by induction of *PDK4* and *RGS1*. Fibroblasts and endothelial cells also shifted towards inflammatory and remodelling-associated states, with enrichment of “Interferon alpha” and “Interferon gamma response”, “Interleukin 10 signalling and “Cytokine receptors bind chemokines” signatures. Fibroblasts showed induction of *FOS*, *DUSP1* and *RGS1*, whereas endothelial cells showed induction of *CCN1*, *SERPINE1*, *APOLD1* and *KLF6*, genes associated with vascular activation and extracellular matrix remodelling.

T cells, B cells and mast cells showed their main signal at the pathway level rather than through large DEG burdens. T cells displayed strong enrichment of both “Interferon alpha” and “Interferon gamma response”, together with more modest enrichment of “IL6-JAK-STAT3 signalling”, “Inflammatory response”, “Allograft rejection” and “TNFα signalling via NF-κB”, consistent with coordinated activation of the lymphoid compartment. B cells and mast cells, despite their low abundance, showed a more restricted response dominated by “Interferon gamma response” enrichment, together with “Inflammatory response” and “Antigen processing cross presentation”. Thus, denosumab-associated immune activation extended across multiple immune populations. Plasmablast and perivascular signals remained limited and were therefore not interpreted further (Fig. 2F; Supplementary Table S4).

Together, this cohort provided a biological informative, clinically diverse framework that revealed at high spatial resolution the multicompartment transcriptional remodelling driven by denosumab within the luminal breast cancer TME.

### Patient-level differential expression and pathway analyses reveal convergent rewiring after denosumab

To assess how these changes varied across individual tumours, we performed differential expression and GSEA separately for each patient and annotated cell population using Hallmark, Reactome and KEGG signatures (Extended Data Fig. 4D; Extended Data Fig. 5; Supplementary Table S5). In line with their cell composition and better sequencing quality, ptx 6, ptx 38 and ptx 61, and to a lesser extent ptx 4, contributed most of the TME signal, whereas changes in ptx 41 and ptx 67 were mainly restricted to tumour cells and fibroblasts. Despite this interpatient variability, the most recurrent post-treatment features were again the enrichment of “TNFα signalling via NF-κB”, “Interferon alpha” and “Interferon gamma response” and several inflammatory and immune programmes, present in all patients except ptx 41, whose tumour sample corresponds to a solid papillary carcinoma lacking stromal TILs (Supplementary Table S3). Among all patients, ptx 38 showed the clearest interferon-driven programme across all cell compartments.

In addition to inflammatory remodelling, denosumab was associated with altered tumour cell state. Proliferation-related programmes were reduced in tumour cells from most patients (notably ptx 4, ptx 38 and ptx 67 and to a lesser extent ptx 41 and ptx 61) (Extended data Fig. 6A), in line with full transcriptome results from the whole cohort showing reduction in “E2F targets”, “G2M checkpoint” (Extended Data Figure 5; Figure 1C; Supplementary Table S2 and S5). Consistently, cell-cycle scoring showed an increased G1 fraction in several cases, supporting a shift towards a less proliferative transcriptional state (Extended Data Fig. 6A), despite the absence of a uniform decrease in Ki67 in the whole patient cohort. In line with this, PAM50 analysis revealed a luminal B-to-luminal A switch in ptx 38 and ptx 67 (Supplementary Table S3), whereas epithelial-to-mesenchymal (EMT)-related programmes were reduced in ptx 6, ptx 61 and ptx 41, consistent with treatment-associated cell-state plasticity.

Thus, despite differences in transcriptional architecture across patients, denosumab-treated tumours converged on coordinated immune rewiring of tumour cells and their microenvironment, as evidenced by pseudobulk and patient-level analyses.

### Bulk-derived inflammatory programmes are spatially distributed across tumour and microenvironmental compartments

Because spatial transcriptomics was performed in a limited number of tumours, we next asked whether these cases captured denosumab-associated transcriptional programmes identified in the broader cohort of 31 luminal tumours from the D-BIOMARK trial. To address this, we projected denosumab-modulated bulk RNA-Seq signatures (selecting leading edge genes from the denosumab arm) into the Visium HD data and assessed their distribution across annotated tumour and TME compartments (see Methods). We focused on samples from patients 6, 38 and 61 because they showed sufficient non-tumoural representation to resolve compartment-specific responses.

We projected selected signatures onto the spatial data, recapitulating the main features significantly altered by ssGSEA in the experimental arm but not in controls (Extended Data Fig. 1C; Supplementary Table S2). Projection onto the Visium HD sections revealed consistent post-treatment spatial changes in ten denosumab-driven programs (see filtering criteria at Methods) including “Hypoxia”, “Inflammatory response”, “Interferon gamma response”, “JAK-STAT signalling” and “Toll-like receptor signalling” (Fig. 2G; Supplementary Table S6).

Importantly, immune programmes were not confined to immune compartments but were also detectable in malignant cells, fibroblasts and endothelial cells. Denosumab-induced enrichment of the bulk “interferon gamma response” signature was particularly prominent in ptx38, where it was observed across tumour, myeloid, fibroblast, and lymphoid populations. Nevertheless, the strongest increase in signature activity was consistently detected in T cells across all three patients (Fig. 2G; Supplementary Table S6). Consistent with compartment-specific biology, “Toll-like receptor signalling pathway” showed its most consistent and robust increase in myeloid cells (Fig. 2G), whereas “Hypoxia” was driven predominantly by endothelial cells (Supplementary Table S6). Notably, non-immune populations also contributed substantially: endothelial cells contributed to the increase in “Inflammatory response”, and “JAK-STAT signalling”, whereas fibroblasts showed a consistent increase in “Interleukin-17 signalling” across the three cases (Supplementary Table S6).

Together, these data indicate that denosumab-associated inflammatory programmes identified by bulk RNA-seq in the 31 denosumab treated patients are spatially embedded within both tumour and stromal compartments. Their recurrence across patients reinforces the value of the spatial analysis for assessing general denosumab-associated effects and supports a multicompartment inflammatory response to denosumab in luminal breast cancer.

### The TME spatially shifts towards an immune enriched milieu after denosumab treatment

Next, we evaluated whether denosumab treatment was associated with changes in the spatial gene distribution. An unbiased spatial generalised additive model (GAM) analysis was performed to systematically identify genes displaying distance-dependent expression patterns relative to tumour regions, without prior selection of predefined pathways or cell states. Functional enrichment of genes significantly enriched in the tumour vicinity (mean derivative >0 and p_adj < 0.001, Supplementary table S7) consistently identified the Hallmark “Epithelial-mesenchymal transition” signature, enriched in collagens and ECM genes, as the top distance-dependent pathway in biopsy samples. This signature showed a clear decrease after denosumab treatment, as reflected by lower odds ratio (OR) and significance. In contrast, immune pathways including “Interferon gamma response” and “TNF signalling” were absent or poorly represented in biopsy samples but became strongly enriched across the tumour boundary after denosumab treatment, with higher OR and significance, supporting enhanced immune infiltration and activation after treatment (Fig. 3A; Supplementary Table S7). This analysis revealed spatial remodelling of the TME following denosumab treatment, characterised by a transition from predominantly fibroblast-associated transcriptional programmes toward an immune-infiltrated reactive niche (Fig. 3A; Supplementary Table S7).

**Figure 3.**
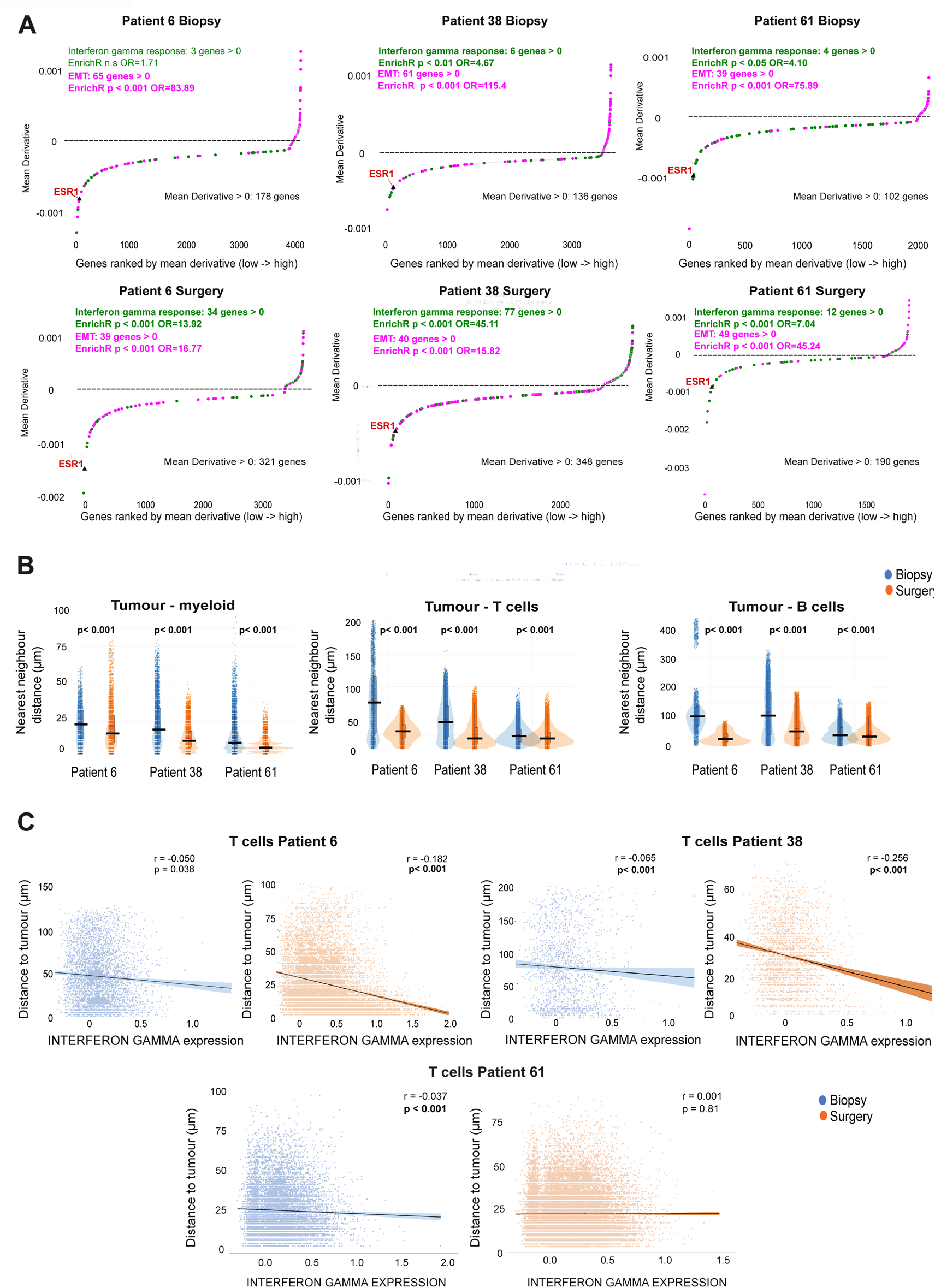
Denosumab increases tumour-immune proximity and interferon-linked activity. **A.** Generalized additive model (GAM) analysis of gene expression changes as a function of distance from tumour regions in biopsy and surgery Visium HD samples from patients 6, 38 and 61. Genes are ranked by mean derivative along the tumour-distance axis. Positive derivatives indicate genes increasing with distance from tumour; negative derivatives indicate genes enriched closer to tumour. Genes belonging to Hallmark Interferon-gamma response and Hallmark epithelial-mesenchymal transition (EMT) signatures are highlighted. The number of genes from each signature with mean derivative > 0 is indicated. Odds ratios and p-values from EnrichR analysis are indicated. Only genes with a significant distance-associated GAM term (p < 0.001) are plotted. Note that the odds ratio and significance for interferon-gamma response increase after treatment, whereas the opposite pattern is observed for EMT. **B**. Nearest-neighbour distances from tumour spots to myeloid cells, T cells and B cells in immune-informative Visium HD cases. Violin plots compare biopsy and surgery samples for patients 6, 38 and 61; p-values are shown. **C**. Relationship between the expression of selected denosumab-associated interferon-gamma signature genes (based on leading-edge genes) in T cells and distance from T cells to tumour regions in patients 6, 38 and 61. Points represent Visium HD T cell spots per patient and condition; lines show linear regression with 95% confidence intervals. Correlation coefficients (r) and p-values are shown for biopsy and surgery. See also Extended Data Figure 6 and Supplementary Table S7

Based on GAM results, we hypothesised that this multicompartment transcriptional remodelling driven by denosumab may be accompanied by measurable changes in the spatial organization of immune and tumour cells. Using tumour spots as reference regions of interest, we quantified distances to T cells, B cells and myeloid cells in patients 6, 38 and 61. Post-treatment samples showed shorter distances between tumour and immune populations, including myeloid cells, T cells and B cells, consistent with increased immune–tumour spatial proximity after denosumab treatment in the three patients (Fig. 3B). Thus, denosumab was associated not only with transcriptional remodelling, but also with measurable reorganization of immune-cell localization within tumour tissue.

To test whether this increased proximity was linked to immune activation, we examined the relationship between tumour distance and “Interferon gamma response” scores within immune populations. In myeloid, T and B cell populations, shorter distance to tumour spots correlated with higher “Interferon gamma response” scores, and this relationship was strengthened after denosumab treatment in patients 6 and 38 (Fig. 3C, Extended Data Fig. 6B-C). In parallel, cell-cycle scoring indicated increased proliferative activity, reflected by an increased S/G2M fraction in T cells from surgical samples compared with biopsies (Extended Data Fig. 6D), consistent with activation of adaptive immune populations. Together, these findings suggest that denosumab enhances immune–tumour crosstalk by promoting immune cell repositioning towards tumour regions, thereby reinforcing the inflammatory and immune-activation programmes identified by bulk and spatial transcriptomic analyses.

### Cell–cell communication analysis identifies reduced tumour-centred signalling and increased T cell-directed communication after denosumab

Given the immunomodulatory effects of denosumab observed in the bulk and spatial analyses, together with the increased immune–tumour proximity after treatment, we next sought to identify candidate mediators of intercellular communication within the TME. To infer spatial cell–cell communication from Visium HD data, we applied CellChat using a 50-μm neighbourhood interaction scale. Communication involving immune populations was again restricted to high quiality immune-informative cases, namely patients 6, 38 and 61.

At the global network level, the most consistent treatment-associated change was a reduction in tumour-centred interactions together with an increase in T cell-associated communication (Fig. 4A-B; Supplementary Table S8). Across the three cases, both outgoing and incoming tumour communication decreased after treatment, whereas communication converging on T cells increased, including signals from tumour, fibroblast, endothelial, myeloid and B cell compartments. This shift was most evident in patients 38 and 61, and was also detectable in ptx 6, although with a weaker T cell output programme. Thus, denosumab appeared to reorient the communication network away from tumour-autonomous signalling and towards a more immune-engaged state.

**Figure 4.**
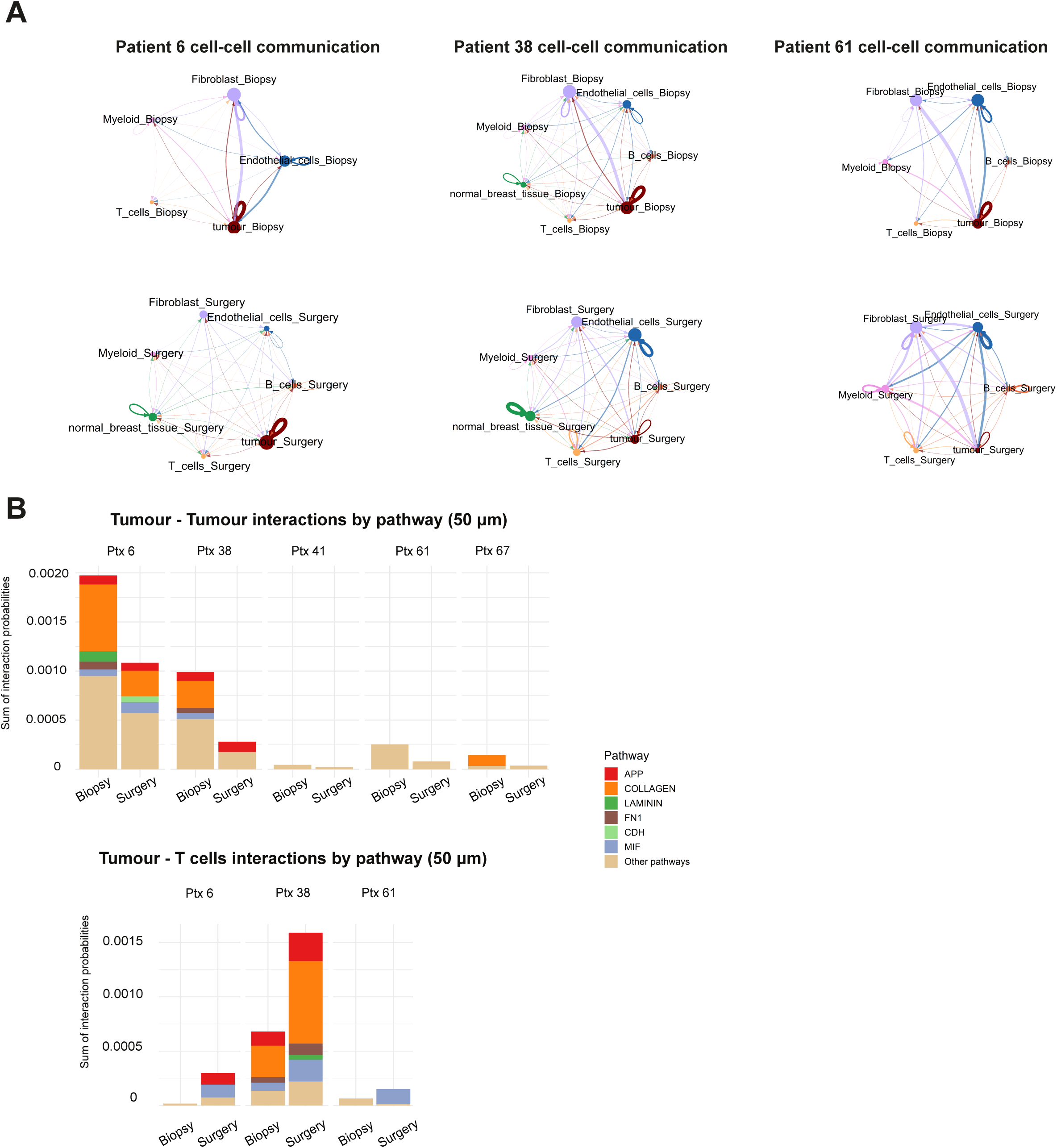
Cell-cell communication analysis identifies reduced tumour-centred signalling and increased T cell-directed communication after denosumab. **A.** CellChat-inferred communication networks in biopsy and surgery samples from immune-informative patients 6, 38 and 61, using a 50-μm neighbourhood interaction scale. Nodes represent annotated cell populations and edges represent inferred interaction strength. **B**. Pathway-level CellChat interaction probabilities for tumour-tumour and tumour-T cell communication across patients. Stacked bars show summed interaction probabilities for each signalling pathway before and after treatment; pathway colours correspond to the CellChat categories shown in the legend. See also Extended data Figure 7 and Supplementary Table S8

This effect was also evident at pathway level. Tumour–tumour communication decreased after treatment in patients 6, 38, 61 and 67, whereas the signal in ptx 41 remained low overall (Fig. 4B). In parallel, tumour–fibroblast interactions were strongly reduced in patients 6 and 38, with weaker changes in patients 61 and 67 (Extended Data Fig. 7). Tumour–myeloid interactions decreased in patients 6 and 38, whereas ptx 61 showed a modest increase. Tumour-endothelial interactions were more variable across patients (Extended Data Fig. 7; Supplementary Table S8). These findings indicate that denosumab does not simply increase communication globally but rather redistributes interaction strength across the network.

At the pathway level, the most reproducible decreases across patients involved ECM- and adhesion-associated signalling, particularly through COLLAGEN, FN1 and LAMININ pathways, which contributed prominently to the reduction in tumour–tumour and tumour–fibroblast communication (Fig. 4B; Extended Data Fig. 7). By contrast, tumour–T cell communication increased through a broader set of inflammatory and immune-associated pathways, including COLLAGEN, LAMININ, APP, and MIF (Fig. 4B). Thus, denosumab attenuated matrix-dominated tumour communication while promoting communication between tumour and T cells, consistent with the increased tumour-T cell proximity observed.

### Tumour clonality inference identifies patient-specific copy number alteration-defined tumour states and treatment-associated shifts in tumour-state abundance

Given the broad transcriptomic and spatial remodelling of the TME induced by denosumab, we next asked whether treatment also altered tumour-state architecture. The tumour compartment contained the largest number of DEGs, suggesting that denosumab may affect tumour-cell transcription directly or indirectly through immune remodelling. To resolve spatial tumour heterogeneity, we applied single-cell variational aneuploidy analysis (SCEVAN) to Visium HD tumour spots using confidently annotated non-malignant reference populations, including immune, endothelial and normal breast cells. Interpretable copy number alterations (CNA)-defined tumour-state maps were obtained for patients 6 and 38 and, more cautiously, for ptx 61. In these cases, SCEVAN resolved discrete tumour subclones with distinct genomic complexity, spatial distributions and transcriptional programmes (Fig. 5A-C). Given the limited non-tumour representation SCEVAN analyses was not feasible for ptx 4, ptx 41 and ptx 67.

**Figure 5.**
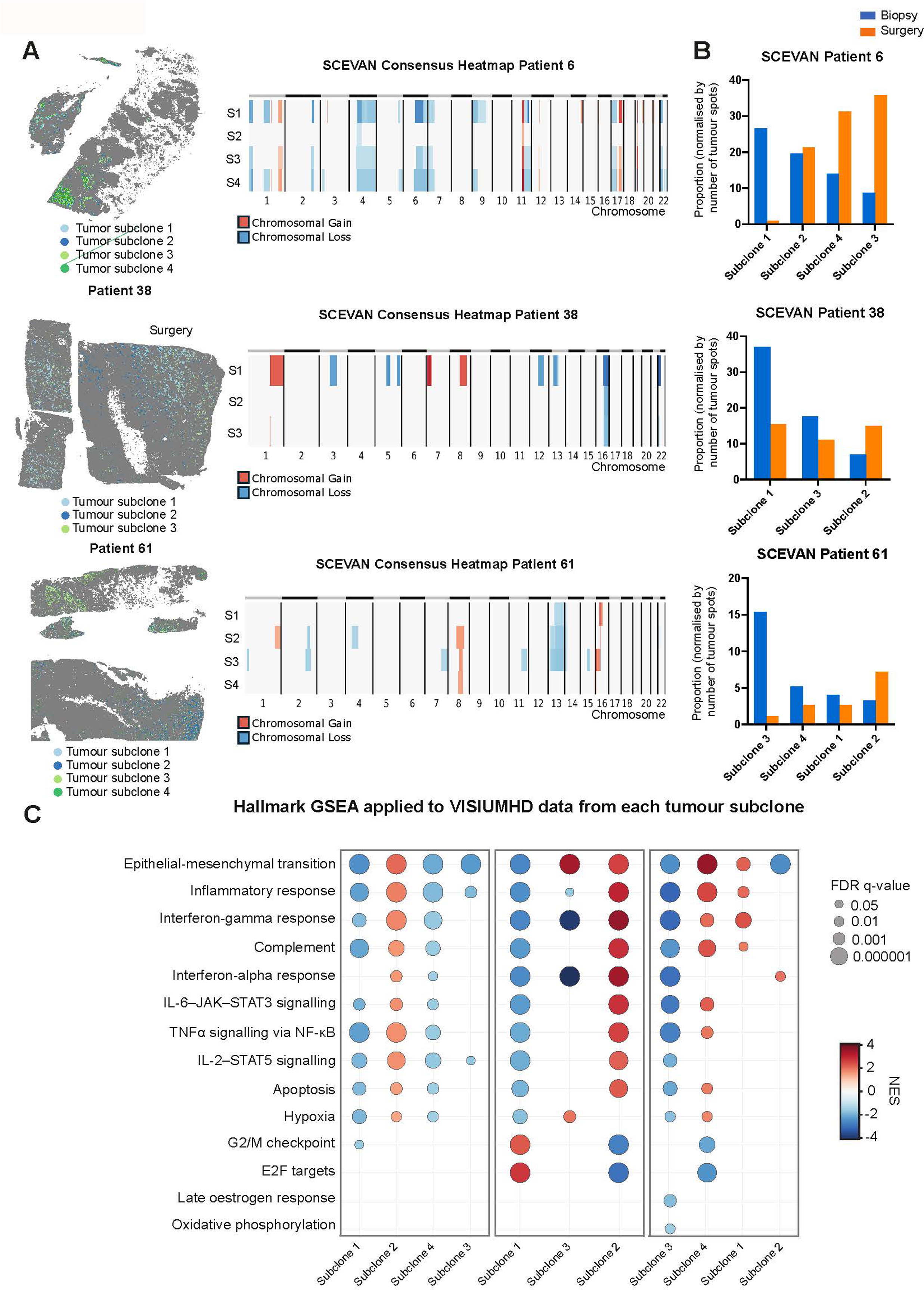
SCEVAN identifies copy number-defined tumour subclones and denosumab-associated shifts in tumour-state abundance. **A.** Spatial distribution of SCEVAN-defined tumour subclones and consensus copy-number heat maps for tumour spots from patients 6, 38 and 61. Rows indicate inferred tumour subclones and columns indicate chromosomes. Colours represent inferred copy-number alterations relative to non-tumour reference populations. **B**. Normalised proportions of SCEVAN-defined subclones in biopsy and surgery samples, calculated relative to the total number of tumour spots per sample. Subclones are ordered from the most abundant to the least abundant subclone in the biopsy sample for each patient. Proportions do not necessarily sum to 100%, because only tumour subclones that passed the filtering criteria for SCEVAN analysis are shown. **C**. GSEA of Visium HD profiles from each SCEVAN-defined tumour subclone. Subclones are ordered from the most abundant to the least abundant subclone in the biopsy sample for each patient, as in **B**. Dot colour indicates NES, and dot size indicates FDR q-value. Positive NES values denote enrichment of the pathway in the corresponding subclone, whereas negative values denote relative depletion. See Supplementary Table S9.

In ptx 6, four tumour subclones were identified, three of which displayed broader CNA burden, including recurrent arm-level losses together with gains on distal 1q and 17q, whereas subclone 2 showed a comparatively simpler profile (Fig. 5A). After treatment, the dominant biopsy state was markedly reduced, whereas subclones 3 and 4 expanded relatively (Fig. 5B). Functional annotation separated the depleted state from the post-treatment-enriched states: the subclone most reduced after denosumab showed weaker enrichment of “Interferon gamma response”, “TNFα signalling via NF-κB”, “IL6-JAK-STAT3 signalling” and related inflammatory programmes, consistent with a more immune-silent phenotype, whereas the retained states were comparatively more inflammatory (Fig. 5C; Supplementary Table S9).

The clearest changes were observed in ptx 38, consistent with the profound activation of tumour immunity observed in this patient. Here, SCEVAN identified three tumour subclones sharing a common CNA backbone, including 1q gain and 16q loss, with additional alterations concentrated in subclone 1, consistent with a more aberrant state (Fig. 5A). In the normalised biopsy *vs.* surgery comparison, subclone 1 was the dominant pretreatment population and decreased after denosumab, with a relative increase in subclone 2 and a reduction in subclone 3 (Fig. 5A-B). GSEA distinguished these states functionally: the depleted subclone was enriched in proliferative pathways (“E2F targets” and “G2M checkpoint”) together with reduction of “Interferon gamma response”, “Inflammatory response” and “Cytokine-cytokine receptor interaction”, whereas the subclones that persisted or expanded after treatment showed the opposite programs (Fig. 5C, Supplementary Table S9). Thus, in ptx 38, denosumab was associated with reduced representation of a proliferative, immune-poor malignant state and relative enrichment of more immune-engaged tumour states.

In ptx 61, SCEVAN again identified multiple CNA-defined tumour states, although the signal was less robust than in ptx 6 and ptx 38. The shared architecture included recurrent 16p gain, 8q gain and losses involving 13q, 15q and 21q, with subclone 3 carrying the most complex profile (Fig. 5A). This subclone was also the dominant pretreatment population and decreased after denosumab, whereas subclone 2 increased relatively and subclones 1 and 4 remained minor populations (Fig. 5B). GSEA again separated the depleted dominant clone from the residual states, with the former showing lower enrichment of immune and inflammatory programmes (Fig. 5C, Supplementary Table S9). Although this case should be interpreted more cautiously, it was consistent with the pattern observed in patients 38 and 6, in which denosumab was associated with reduced representation of the dominant, more CNA-complex and less immune-associated malignant state.

Taken together, these analyses identify a convergent treatment-associated pattern across all three evaluable tumours. In each case, the subclone that decreased after denosumab corresponded to the state with the highest CNA burden and the weakest immune-associated transcriptional features, whereas persistent or expanding clones showed relatively stronger inflammatory or interferon-related programmes. These findings extend the bulk and spatial analyses by suggesting that denosumab does not merely remodel the microenvironment but may also shift the balance of malignant cell states away from genomically complex, proliferative and immune poor subclones.

## DISCUSSION

Despite major advances in cancer immunotherapy, luminal breast cancer remains substantially less responsive to immune-based strategies than more immunogenic breast tumour subtypes (Vonderheide et al., 2017). Although TILs are established predictive and prognostic biomarkers in triple-negative breast cancer, their biological and clinical relevance in luminal disease remains uncertain (Denkert et al., 2018; Schlam et al., 2024). This reflects a challenge: in luminal tumours, immune regulation is unlikely to be captured by immune-cell density alone. Rather, immune accessibility is shaped by the coordinated organization of malignant cells, stromal structures, vascular compartments and immunosuppressive populations. Understanding the mechanisms that regulate this tissue architecture is therefore essential for developing rational immunomodulatory strategies in this setting.

Among these mechanisms, the RANK/RANKL pathway has emerged as a relevant regulator of tumour–immune interactions (Ahern et al., 2018). This study places short-course RANKL blockade within a multicellular model of luminal breast cancer remodelling and provides a framework for understanding how this pathway operates across tumour, stroma and immune compartments.

Building on previous observations from D-BEYOND, the randomised D-BIOMARK trial demonstrated that preoperative denosumab leads to activation of anti-tumour immune programmes within the TME (Gómez-Aleza et al., 2020; Vethencourt et al., 2025). In D-BIOMARK, the inclusion of untreated tumours represents a major strength, as biopsy-to-surgery effects, tissue injury induced by the diagnostic procedure, and temporal variation in immune infiltration can confound the interpretation of treatment-associated changes. However, as the 2:1 randomization resulted in fewer control samples, differences between treated and untreated tumours should be interpreted in relation to effect size, directionality, and biological coherence. The administration of two doses of denosumab followed by surgery within 14–28 days was designed to capture treatment-related biological effects during a period of expected activity (Chen et al., 2018; U.S. Food and Drug Administration, 2025).

Our results show that lymphocyte accumulation alone is an insufficient readout of denosumab activity. Although TIL, CD3+ (and specifically CD8+) frequency also tended to increase in untreated tumours, bulk RNA-Seq revealed a broader and coordinated biological response in treated samples, characterised by enrichment in inflammatory, innate and adaptive immune pathways, including interferon and TNF signalling, together with attenuation of tumour-associated proliferative programmes. Moreover, deconvolution analysis suggested a coordinated increase in lymphocyte and monocytic signatures, together with a reduction in immunosuppressive macrophage-(and Treg-) associated signatures after (Peranzoni et al., 2018) denosumab treatment. Rather than reflecting simple changes in immune cell abundance, these findings suggest functional remodelling of the immune compartment. Spatial analyses reinforced that myeloid and lymphoid remodelling is a central correlate of the denosumab response, although additional effects in tumour cells, fibroblasts and endothelial cells are likely.

The greater magnitude and diversity of transcriptomic changes observed after denosumab exposure exceeded with untreated tumours over the same biopsy-to-surgery interval further support a treatment-associated biological effect. These findings are consistent with previous observations from D-BEYOND (Gómez-Aleza et al., 2020) which suggested activation of anti-tumour immune programmes following RANKL blockade and are further strengthened here by the controlled design of D-BIOMARK.

Beyond the local tumour microenvironment, denosumab exposure was also associated with systemic changes in the overall patient cohort. In paired serum samples, denosumab exposure was associated with reduced TNFRSF9 (also known as 4-1BB) and CD244 levels, alongside increased TNF and ARG1, proteins with important roles in immune regulation (Müller, 2023). TNFRSF9 has additionally been linked to osteoclast biology and bone turnover (Wan et al., 2021; Yang et al., 2008). The increase in circulating TNF is consistent with the inflammatory and T cell activation programmes observed in the TME. Overall, the concordance between local and circulating immune-related changes suggests that RANKL blockade may also be associated with systemic immune modulation.

The strongest bulk-derived denosumab-associated signatures were recapitulated in the spatial analyses and revealed that, rather than acting through a single compartment, RANKL blockade was associated with coordinated remodelling across malignant, immune, fibroblast and endothelial populations. Beyond immune cell composition, the reproducible endothelial signal and reduced matrix-associated communication suggest that denosumab may alter the structural context in which immune responses occur. Fibroblasts, endothelial cells and extracellular matrix organisation have emerged as active regulators of immune accessibility and treatment response, consistent with observations linking RANK pathway modulation to changes in stromal and collagen-associated programmes (Simões et al., 2025). Accordingly, stromal remodelling in our study was accompanied by signals compatible with increased immune accessibility and immune activation. This is particularly relevant in luminal breast cancer, where tissue architecture and spatial organisation are increasingly recognised as key determinants of immune regulation and treatment response (Jackson et al., 2020). Recent spatial transcriptomic work has shown that cancer hallmarks are spatially organised within tumours and that their distribution reflects tumour ecological dynamics and regional heterogeneity (Sibai et al., 2025). Therefore, analyses accounting for tissue area, tumour area, stromal content and immune cell representation are important to distinguish treatment-associated remodelling from sampling bias.

The reduced distance between tumour and T cells, together with features of T cell activation, including increased T cell proliferation and interferon/TNF-response programmes, is compatible with enhanced tumour–immune engagement after denosumab exposure. Although exploratory, the SCEVAN analysis raises the possibility that the reduction of the most CNA-complex and least immune-associated clones, may reflect immune selection or immunoediting. This observation is particularly interesting in light of previous reports linking high copy-number burden with reduced immune recognition and resistance to immune checkpoint blockade across cancers (Asleh & Ouellette, 2024). However, this interpretation should remain cautious, as SCEVAN infers broad CNA-defined states from transcriptomic data rather than directly measuring focal genomic alterations.

The spatial transcriptomic cohort was intentionally focused on biologically informative paired samples, enriched for luminal B-like tumours, to resolve the cellular and spatial architecture of the denosumab-associated immune response. These analyses were supported by paired bulk RNA-Seq from the broader cohort, which showed concordant post-treatment enrichment of inflammatory and immune-regulatory programmes. Agreement across platforms supports the biological representativeness of the spatial data, although larger outcome-linked studies will be required to determine the clinical implications of these findings.

Despite major concordant responses, heterogeneity in the magnitude and pattern of response across tumours remained evident. The stronger spatial responses were observed in the three cases showing a greater than 10% increase in TILs, whereas ptx 41 remained discordant, likely reflecting both biological and technical factors, including tumour-dominant architecture, lack of immune representation and the fact that this tumour was histologically catalogued as a solid papillary breast carcinoma. More broadly, these data argue that menopausal status, luminal A/B biology, immune composition, histological architecture, tumour cellularity and tissue immune content may all influence the magnitude or detectability of response to denosumab.

This context dependency may also help explain the heterogeneous results observed across previous clinical studies of adjuvant denosumab in early breast cancer, including ABCSG-18 and D-CARE, which differed substantially in patient population and trial design (Coleman et al., 2020; Gnant et al., 2019), as well as GeparX, in which denosumab was administered together with neoadjuvant chemotherapy but did not translate into improved clinical outcomes (Blohmer et al., 2022).

Beyond breast cancer, clinical trials such as CHARLI, KeyPAD and POPCORN have shown that combining denosumab with immune checkpoint inhibitors appears feasible and may enhance anti-tumour activity without introducing major additional toxicity (Ahern et al., 2019, 2026; Gedye et al., 2023; Lau et al., 2023). Whether the transcriptional and spatial alterations observed in D-BIOMARK translate into enhanced clinical benefit remains unknown. However, combining denosumab with immune checkpoint inhibitors represent a biologically distinct strategy that deserves further investigation.

Overall, our findings support a model in which RANKL blockade remodels luminal breast cancer through coordinated changes across tumour, immune and stromal compartments. This framework supports future biomarker-guided studies evaluating denosumab as part of immunomodulatory strategies in luminal breast cancer.

## METHODOLOGY

### Study Design

The D-BIOMARK clinical trial (NCT03691311) is a window-of-opportunity, randomised study sponsored by the Institut Català d’Oncologia. A total of 60 patients were enrolled between August 2018 and May 2021. For this publication, only cases of estrogen receptor (ER) positive breast cancer expressing or not progesterone receptor (PR) (luminal tumours, n = 48) were included. Patients with early-stage HER2-negative breast cancer who were candidates for tumour excision as the first therapeutic approach were selected. Participants were randomised in a 2:1 ratio to either the experimental group, which received two subcutaneous doses of denosumab (120 mg each, separated by 7 days), or the control group, which received no treatment. Informed consent was obtained from all patients before entering the study, and the study protocol was approved by the Research Ethics Committee of the Hospital Universitari De Bellvitge (protocol code PR035/21), adhering to ethical standards outlined in the Declaration of Helsinki (1964). The study adhered to Good Clinical Practice (GCP) guidelines as per the International Conference on Harmonization (ICH) Tripartite Guidelines 1996 and complied with regulations on patient data access and data protection (Ley Orgánica 15/1999). The tumour biopsies analysed were diagnostic core-needle biopsy (pre-treatment) and surgical specimen (post-treatment, 2-4 weeks after inclusion). Serum samples were collected at these two time points (biopsy and surgery). Follow-up visits for safety evaluation occurred one month and six months post-surgery.

### Immunohistochemistry in D-BIOMARK tumour samples

The evaluation of conventional BC markers, including ER and PR, was performed in the Pathology Department of the Hospital Universitari de Bellvitge. The status of ER and PR was defined according to the guidelines of the American Society of Clinical Oncology and the College of American Pathologists (ASCO-CAP 2010), whereas HER2 status was assessed and interpreted according to the updated ASCO/CAP guidelines for HER2 testing in breast cancer (Hammond et al., 2010). The histological grade was evaluated following the Nottingham scoring system. TILs were evaluated on H&E-stained slides using standardised methodology (Salgado et al., 2015). The jackischsurrogate subtypes of BC were defined according to the St. Gallen consensus 2015 using immunohistochemically surrogates as follows: Luminal A: ER and/or PR(+), HER2(−), Ki67 < 20%; Luminal B: ER and/or PR(+), HER2(−), Ki67 ≥ 20% (Jackisch et al., 2015). RANK and RANKL were assessed using 4 μm paraffin tissue sections, with IHC staining performed at NeoGenomics Laboratories (California, USA). Pathological evaluations, including tumour cellularity and tumour-infiltrating lymphocytes, were conducted by two blinded pathologists. Immune cell infiltration was analysed via IHC, staining for CD4, CD8, CD20, CD68, MUM1, CD4/FoxP3, PD1, and PDL1. The quantification of all parameters, except PDL1, was performed using QuPath® software (a bioimaging analysis platform). PD-L1 staining intensity was assessed in both tumour and stromal cells and categorised into four levels: level 0 (no staining), level 1 (weak staining), level 2 (moderate staining), and level 3 (strong staining). For each case, three representative fields were randomly selected for evaluation. The H-score, a semi-quantitative measure of PD-L1 expression, was then calculated using the following formula: H-score (range 0–300) = [0 × (% of unstained tumour cells)] + [1 × (% of weakly stained tumour cells)] + [2 × (% of moderately stained tumour cells)] + [3 × (% of strongly stained tumour cells)].

### Gene expression analysis of D-BIOMARK tumour samples

RNA was extracted from O.C.T.™ samples at baseline and at the time of surgery. Before extraction, a section of the sample was stained with H&E to evaluate the percentage of tumour content. A tumour cellularity of at least 30% was deemed necessary for RNA extraction. Total RNA was isolated from tumour specimens using the Maxwell^®^ RSC simplyRNA Tissue Kit (AS1340 Promega), following the manufacturer’s instructions. A total of 37 RNA paired samples, in the range of 1-70 ng, with RIN in the range of 2.5-8.0, were processed into cDNA sequencing directional libraries with the “QuantSeq 3‘mRNA-Seq Library Prep Kit (FWD) for Illumina” (Lexogen, Cat. No. 015). A Unique Molecular Identifier (UMI) Second Strand Synthesis module was used to address bias correction. Libraries were completed by PCR and sequenced on an Illumina NextSeq instrument by following manufacturer’s protocols.

The data analysis pipelines included UMI extraction (UMI-tools v1.1.0) (Smith et al., 2017), Trimming (BBMap v38.93) (Bushnell, 2014), Alignment (STAR V2.7.8a) to the GRCh38 reference genome (Dobin et al., 2013), UMI collapsing (UMI-tools dedup), Gene counting (HTSeq-count v1.99.2) (Anders et al., 2015) and Differential gene expression analysis (DESeq2 v1.34.0) (Love et al., 2014).

Gene set enrichment analysis (GSEA) pre-ranked analysis was performed using GSEA (version 4.2.2) (Subramanian et al., 2005). The ranked lists used for the analysis were derived from the results of the differential expression analysis. To generate these ranked lists, Ensembl IDs were converted to gene symbols with R bioconductor biomaRt (version 2.50.0) (Durinck et al., 2009) with the hsapiens gene ensembl dataset (Cunningham et al., 2022). For each gene symbol, the maximum (in absolute value) of the test statistical values was selected. Non-coding RNAs were then excluded, and the list was sorted by the test statistical value. The parameters used for the GSEA analysis were the default settings, with the following exceptions: -collapse false, -chip gseaftp.broadinstitute.org://pub/gsea/annotations/GENESYMBOL.chip, -plot top x 100, and -set min 10. ssGSEA was performed using the GenePattern platform (Broad Institute) to calculate enrichment scores for predefined gene signatures in each bulk RNA-Seq sample. The resulting ssGSEA scores were exported for downstream analyses. For patients with paired biopsy and surgical samples, changes in pathway activity were calculated as Δ score (surgery − biopsy).

Differences in Δ scores between groups (Control vs Experimental) were assessed using two-sided Welch’s t-tests, and effect sizes were estimated using Cohen’s d. Longitudinal changes within each group were evaluated using paired two-sided t-tests. Pathways with nominal p < 0.05 were considered significant. To further refine the immune cell subsets, present in each sample, CIBERSORTx (Newman et al., 2019) software was used. mRNA sequencing data were handled anonymously.

#### Serum proteomic analysis in D-BIOMARK serum samples

Proteomic analysis was performed using the Olink platform, a specialised service that employs advanced technologies for protein analysis and biomarker discovery. The Immuno-Oncology panel, consisting of 96 proteins related to immune responses in neoplastic diseases, was used (https://olink.com/products/olink-target-96). Olink’s Proximity Extension Assay (PEA) technology was employed, where antibody-based proximity triggers DNA extension, followed by quantification via qPCR or NGS. Data was analysed using relative quantification with normalised protein expression (NPX) units to facilitate comparative analysis.

### Biomaterials preparation for Visium HD

A certified pathologist selected a region of interest, excluding necrotic and fibrotic areas, for the preparation of specimen blocks. RNA quality of the tissue block was assessed by calculating the percentage of total RNA fragments > 200 nucleotides (DV200) as recommended in the Visium HD FFPE Tissue Protocol (10x Genomics, document CG000684). All tissue samples used for the Visium HD Spatial Gene Expression had a DV200 of > 30%, in accordance with this protocol. The Histopathology Core Unit of the CNIO performed tissue sectioning and sample preparation. Biopsy and surgical specimens from the same patient were embedded in the same paraffin block and processed following the Visium HD FFPE Tissue Protocol (10x Genomics, document CG000684).

### Visium HD Spatial Transcriptomics libraries preparation

Hematoxylin and eosin (H&E) staining and imaging were performed according to the Visium HD FFPE Tissue Preparation Handbook (CG000684). Samples were then processed and sequenced following the Visium HD Spatial Gene Expression Reagent Kits User Guide (CG000685) using Visium HD v1-FFPE chemistry. Eluted probes were converted into sequencing libraries by pre-amplification and post-sample index PCR as recommended by the manufacturer. Libraries were sequenced on an Illumina NovaSeq X according to the manufacturer’s instructions.

### Visium HD gene expression quantification

Raw sequencing files were mapped to Visium Human Transcriptome Probe Set v2.0 and GRCh38-2020-A transcriptome and gene expression quantification was performed by spaceranger count command (spaceranger-3.0.0 pipeline version). Output matrices were generated for 2, 8 and 16um resolution bins and used for downstream analyses using Seurat package (v.5.3.1) (Hao et al., 2024).

### Cell-type annotation

Tumour cells and normal breast epithelium were manually annotated by a certified pathologist from the Spanish National Cancer Research Center (CNIO) using 8-μm binned cloupe file in Loupe Browser (10x Genomics Loupe Browser v8.0.0) software. These annotated barcoded spots were converted to 16-μm bin resolution using loupeR package (v.1.1.4) (Siegel, 2025)and then exported as CSV files for downstream analysis. Stromal and tumour microenvironment (TME) cell composition was elucidated using scRNA-Seq data for cell-type deconvolution. Briefly, two datasets from human breast cancer scRNA-Seq and normal breast single-nucleus RNA-sequencing (snRNA-Seq) (Kumar et al., 2023; Pal et al., 2021) were chosen for cell type deconvolution using Robust Cell Type Decomposition (RCTD) approach (spacexr R package v.2.2.1) (Cable et al., 2022). For Pal et al. dataset, only the ER+ breast tumours from GSE161529 series were kept. This dataset allowed us the annotation of the following cell types: B cells, T cells, endothelial cells, myeloid cells, plasmablasts, and stromal, which correspond mainly to fibroblast. To identify cell types that are underrepresented in droplet-based scRNA-Seq datasets due mainly to size constraints (adipocytes and mast cells), we leverage on Kumar et al. snRNA-Seq dataset from normal breast tissue (GSE234817). Deconvolution from this dataset allowed the assignment of the following identities: lymphatic cells, adipocytes, fibroblast, perivascular cells, and mast cells. Final annotation was then built by combining the pathologist assignment for the tumour and epithelial compartment with the RCTD-deconvoluted maximum class score identity for the stromal and TME spots. For those spots deconvoluted as epithelial or tumour cells that were not annotated by the pathologist, we created normal breast mixed and tumour mixed identities, respectively. These spots mainly correspond to regions in the tumour borders and surrounding normal epithelium, including also stromal tissue.

### Individual analysis

Each capture area was analysed individually using 16um resolution matrices using Seurat (v5.3.1). Final annotation described in the section above was used for filtering low counts spots and incorporated as metadata. Downstream analysis included all standard Seurat analytical steps: normalization, identification of variable features, scaling, clustering, and dimensionality reduction (DeTomaso et al., 2019). GSEA by patient was carried out through fgsea R package (v.1.36.2) using ranked gene expression list as input provided by Seurat’s FindMarkers function and the same gene sets. To assess treatment-associated changes in proliferation, Seurat’s CellCycleScoring function was used to determine the cell cycle phase scores for each cell population. Then, biopsy and surgery distributions of the proliferative state (S+G2M) were analysed using Fisher’s exact test.

### Pseudobulk analysis

All patients were analysed together by integrating all individual Seurat objects using harmony software (v.1.2.3) using capture area as covariable for correction. Cell population based differential gene expression analysis was performed by aggregating raw gene counts by the final annotation identities for each patient pre and post denosumab treatment, using AggregateExpression command from Seurat. Each cell population raw count matrix was then used for differential expression analysis using the DESeq2 package (v.1.46.0). Gene Set Enrichment Analysis was carried out from median of ratios normalised matrices, using Molecular Signatures Database hallmark gene set collection (h.all.v2025.1.Hs.symbols.gmt), KEGG collection, and Reactome Pathways Gene Set (number of permutations = 1000, seed = 149, and permutation type = gene set).

### Projection of denosumab-modulated genes from the bulk RNA-Seq into Visium HD data

Differential signatures of the experimental arm from bulk RNA-Seq data were projected into the Visium HD data using VISION package (v.3.0.2) (DeTomaso et al., 2019) and signature scores were analysed using a downsampling (DS)-based approach. Filtering criteria include: concordant ΔDS direction across the three patients, significant global DS after false-discovery-rate correction (FDR < 0.05), and a large absolute DS effect size (|effect| > 0.8). Signatures passing this filter were considered high-confidence surgery–biopsy-associated signatures and were used for downstream pathway interpretation. Downsampling values, FDR-adjusted significance values and enrichment results are reported in Supplementary Table 6.

### Distance analysis

STDistance R package (v.0.6.6) [https://github.com/PrinceWang2018/ST Distance] (Wang et al., 2025) was used to calculate the distance to the nearest spot by considering tumour spots as reference and the different cell types as targets. Distribution of these distances between biopsy vs surgery for each patient was evaluated using the Wilcoxon rank sum test adjusted performing the Benjamini-Hochberg procedure. Correlation analysis was carried out using the nearest neighbour distance and the RCTD score of different signatures.

Radial distance analysis was performed using the RadialDistances function of semla R package (v.1.4.0) [https://github.com/ludvigla/semla](Larsson et al., 2023). Spatial distance–dependent gene expression was assessed using generalised additive models (GAMs). For each spatial spot, the radial distance to the nearest tumour region was computed using the Semla framework. Gene expression (log-normalised counts) was then modelled as a smooth function of distance (y ∼ s(r dist tumour)) using the mgcv package in R, employing cubic regression splines with shrinkage to capture potential nonlinear relationships. Analyses were restricted to spots within 0–1000 µm from tumour regions, and genes detected in at least 5% of spots were included. For each gene, the significance of the smooth term was evaluated, and p-values were adjusted for multiple testing using the Benjamini–Hochberg method. To summarize the directionality of spatial effects, the mean first derivative of the fitted smooth curve across the distance range was computed, indicating whether gene expression increased or decreased with distance from the tumour.

### Cell-cell communication inference

Inference of spatially proximal cell-cell communication was performed using the R package ‘CellChat’ (v.2.2.0) (Jin et al., 2025) following the developer’s vignette [https://github.com/jinworks/CellChat], which infers spatially proximal cell-cell communication between interacting cell groups from spatially resolved transcriptomics. In this analysis, a spot size of 16 μm was defined, along with the following parameters: type = “truncatedMean”, trim = 0.1, distance.use = TRUE, scale.distance = 4.3, contact.dependent = TRUE, contact.range = 16 and interaction.range=50. Absolute weight for each pathway was obtained by summing up the probabilities for each ligand-receptor pair.

### Tumour clonal inference

To characterize intra-tumoural heterogeneity and clonality, we applied SCEVAN (v.1.0.3) [https://github.com/AntonioDeFalco/SCEVAN] (De Falco et al., 2023) to Visium HD transcriptomics data, using tumour spots and a reference of diploid cells, which included T cells, B cells, myeloid cells, endothelial cells, and stromal cells. The analysis was performed on samples 6, 38, and 61, as samples 4, 41, and 67 did not pass preprocessing filters. The following parameters of the pipelineCNA function were used: normal cells vector, SUBCLONES = TRUE, beta vega = 0.5, ClonalCN = TRUE, plotTree = TRUE, AdditionalGeneSets = NULL, SCEVANsignatures = TRUE, organism = “human”, ngenes chr = 5, perc genes = 10, FIXED NORMAL CELLS = TRUE). Furthermore, differential gene expression analysis was carried out by Seurat’s FindMarkers function using each subclone against others and the ranked list by log2 fold change was used as input to perform GSEA through fgsea R package (v.1.36.2).

### Statistical analysis

All analyses were performed using R version 4.1.3 (available at www.r-project.org) and GraphPad Prism software version 9.0.1 (GraphPad Software, Boston, MA, USA; www.graphpad.com). For human analysis, values at baseline (biopsy) were compared to those at surgery using paired t-tests for numerical variables and McNemar tests for binary variables. Independent samples t-tests were used to compare differences between groups for continuous variables, with chi-square or Fisher’s exact tests for binary variables when appropriate.

## Data availability

The bulk RNA-Seq and Spatial Transcriptomics data generated in this study have been uploaded to GEO. Bulk RNA-Seq on paired biopsy/surgery tumour samples from the D-BIOMARK clinical trial: GSE291079. Spatial Transcriptomics data are available at GSE335224.

## Code availability

The code associated with the analysis are available at GitHub: (https://github.com/dvalcarcel/RANKL_inhibition_Spatial_Transcriptomics).

## Supplementary Information available for this paper

Extended Data Figures 1-7

Supplementary Tables S1-S9

## Acknowledgements

This work was supported by grants to Eva González-Suárez by the European Research Council (ERC) under the European Union’s Horizon 2020 research and innovation programme ERC-2015-CoG (grant agreement no. 682935), ERC-2021-PoC, SAF2017-86117R and PID2020-116441GB-I00, funded by the Agencia Estatal de Investigacion (AEI/10.13039/501100011033, Ministerio de Ciencia e Innovación) and Caixa Research Health 2023 MYELO-RANK HR23-00361 Grants. FPU23/02826 PhD contract and Residencia de Estudiantes 2025/26 fellowship to M Rodriguez-del-Collado, both by the Ministry of Science, Innovation, and Universities; Rio Hortega contract CM19/00148 to A Vethencourt funded by the Carlos III Health Institute, the Spanish Breast Cancer Research Group GEICAM (Balil-Pelegrì grant to A Vethencourt); PRE2018-086522 PhD contract to A Barranco; and Juan de la Cierva contract IJCI-2017-31564 to EM Trinidad. D-BIOMARK is an Investigator Study sponsored by Institut Català d’Oncologia: Amgen has assisted in the funding. The findings expressed herein are solely those of the authors, and Amgen does not assume any responsibility therefor.

We would like to thank the CNIO Histopathology Core Unit and Genomic Core Unit, particularly Ángeles Rubio. We thank Biobank HUB-ICO-IDIBELL (PT20/00171) integrated into the ISCIII Biobanks and Biomodels Platform, and Xarxa Banc de Tumors de Catalunya (XBTC) for their collaboration, and CERCA programme/Generalitat de Catalunya for institutional support. We thank Francisco X. Real for helpful discussions in the interpretation of the data.

We extend our heartfelt gratitude to all team members involved in the project, encompassing the medical oncology, radiology, pathology, and surgery teams. Special recognition is extended to Ariadna Iserte (ICO) for her exceptional coordination, and to Maria Dolores Mulero and the histology team at Idibell for their assistance with staining procedures. We also thank Juana Moro and Marta Matas for their help with data interpretation and statistical analysis. Finally, we are deeply grateful to the patients and their families who generously participated in this study.

## Institutional Review Board Statement

The study was conducted in accordance with the Declaration of Helsinki and approved by the Institutional Ethics Committee of the “Institut Català d’Oncologia de L’Hospitalet (ICO-L’Hospitalet)” (protocol code **PR035/21**). Informed consent was obtained from all subjects involved in the study.

## Author contributions

Conceptualization: E.G.-S., A.V., C.F. Investigation: M.R.-d-C., A.V., A.B., E.-M.T., E.D., M.J., E.J.C., M.G., C.G., O.D., G.P.-C., M.C., E.P., A.U., A.P., M.T.S.-M., A.G., S.P., C.F. Formal analysis: M.R.-d-C., A.B., A.V., I.S., J.M.-d-V., D.V.-L., G.S.-A., G.G., E.P.-Y., J.N., J.H.C. Data curation: G.S.-A., G.G., E.P.-Y., J.M.-d-V., D.V.-L. Software: J.M.-d-V., D.V.-L. Visualization: M.R.-d-C., A.B., A.V., J.M.-d-V., D.V.-L. Funding acquisition: E.G.-S., A.V., C.F. Supervision: E.G.-S. Writing—original draft: M.R.-d-C., A.V., A.B., J.M.-d-V., D.V-L., E.G.-S. Writing—review & editing: all authors.

---

**Extended Data Figure 1.**
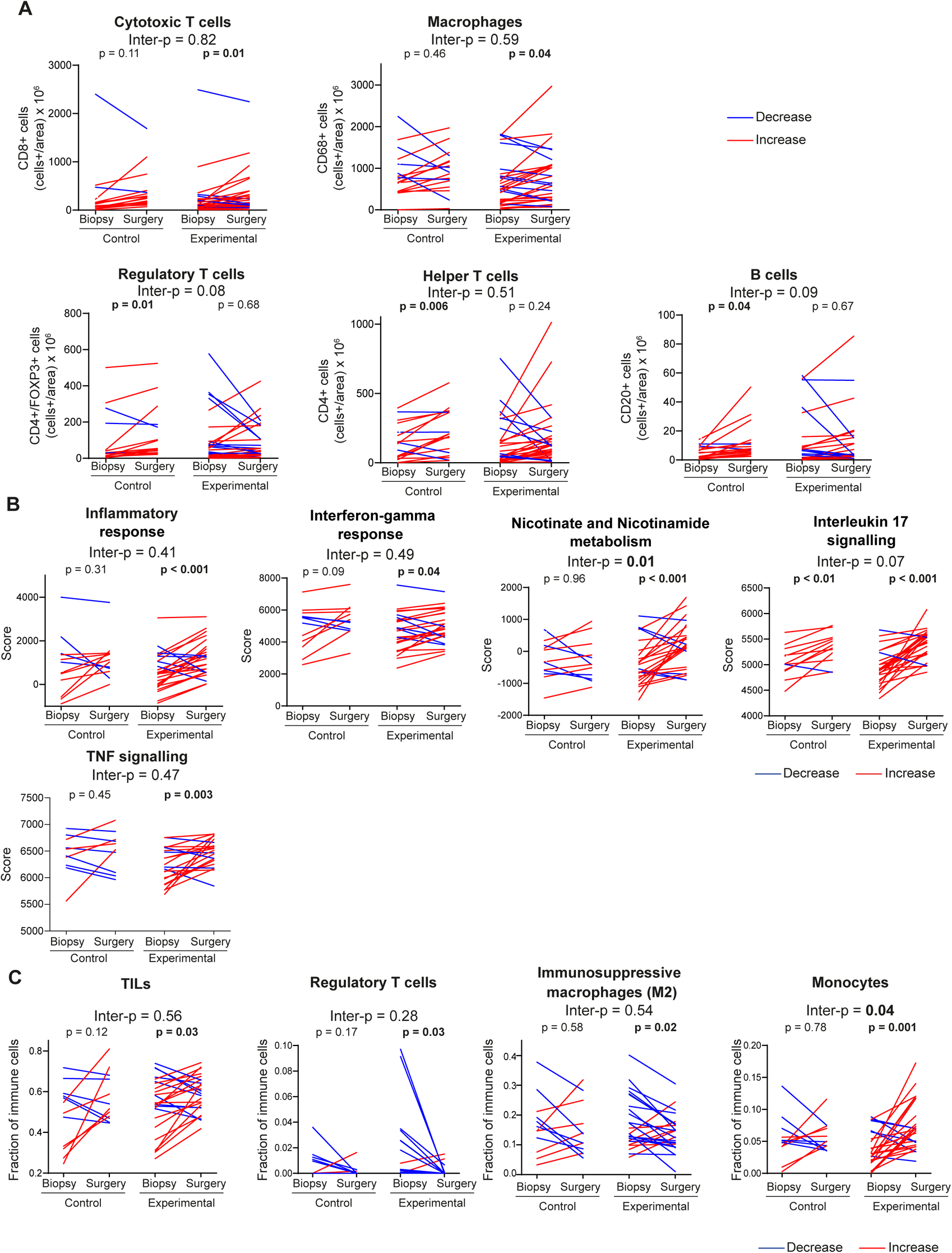
Preoperative denosumab modulates the tumour microenvironment. A. Immunohistochemical quantification of cytotoxic T cells (CD8+), macrophages (CD68+), regulatory T cells (CD4+FOXP3+), T helper cells (CD4+), and B cells (CD20+) in paired biopsy and surgery samples. Values are expressed as positive cells per tissue area (×10^6^). B. Single-sample pathway scores for selected bulk RNA-Seq signatures in paired biopsy and surgery samples. C. CIBERSORT-inferred immune cell fractions in paired biopsy and surgery samples, showing total TILs, regulatory T cells, immunosuppressive (M2-like) macrophages and monocytes. A-C. Red and blue lines denote increases and decreases from biopsy to surgery, respectively. P-values above pairs compare biopsy with surgery within each arm; inter-p values compare surgery-biopsy changes between arms.

**Extended Data Figure 2.**
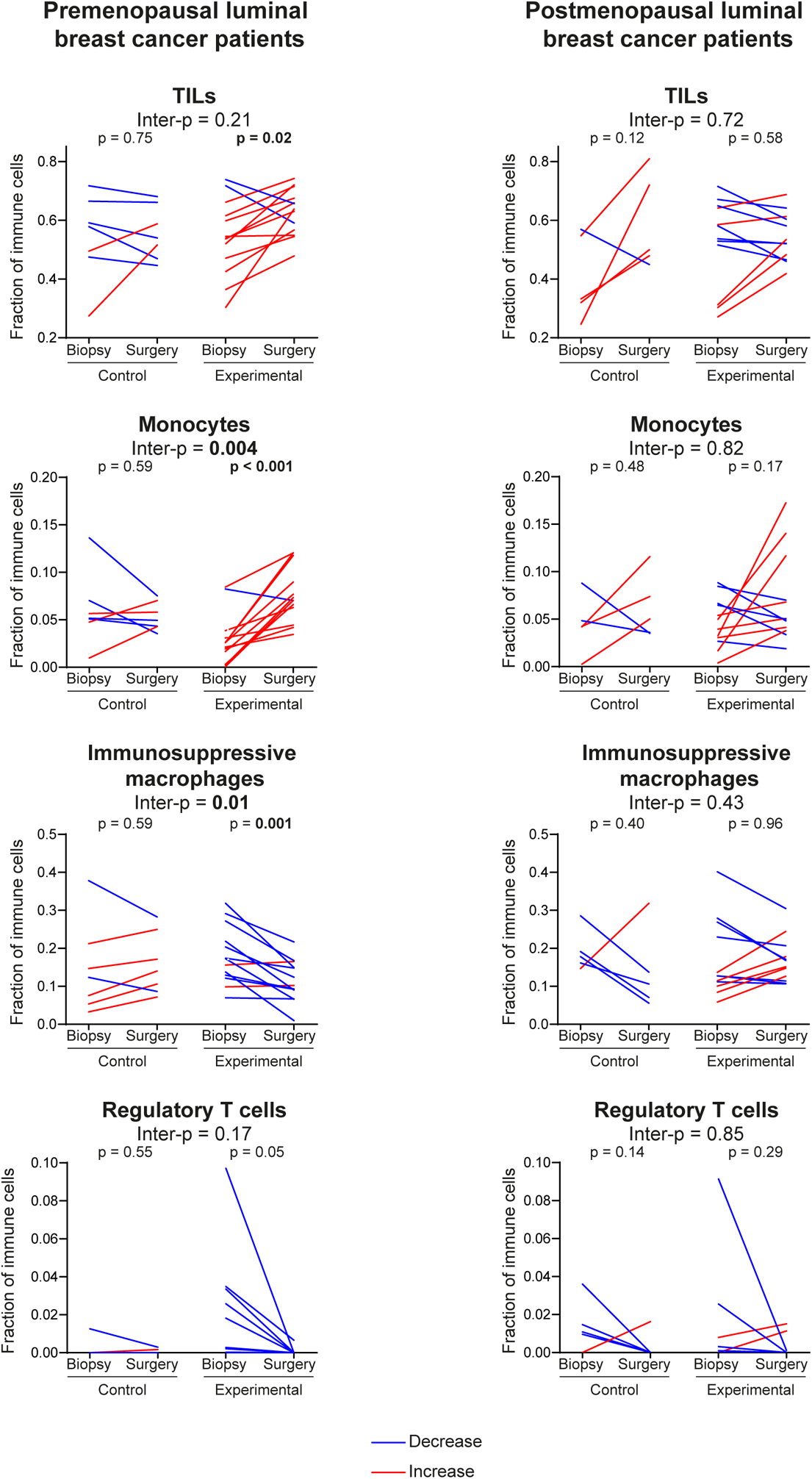
Denosumab immunomodulatory effects are more pronounced in premenopausal luminal breast cancer patients. CIBERSORT-inferred fractions of TILs, monocytes, immunosuppressive macrophages and regulatory T cells in paired biopsy and surgery samples from premenopausal (control n = 7; denosumab n = 13) and postmenopausal (control n = 5; denosumab n = 12) patients with early-stage luminal breast cancer in D-BIOMARK. Values are proportions from 0 to 1. Red and blue lines denote increases and decreases from biopsy to surgery, respectively. P values above paired plots are from paired t-tests within each arm; inter-p values compare surgery-biopsy changes between denosumab and control arms by unpaired t-test.

**Extended Data Figure 3.**
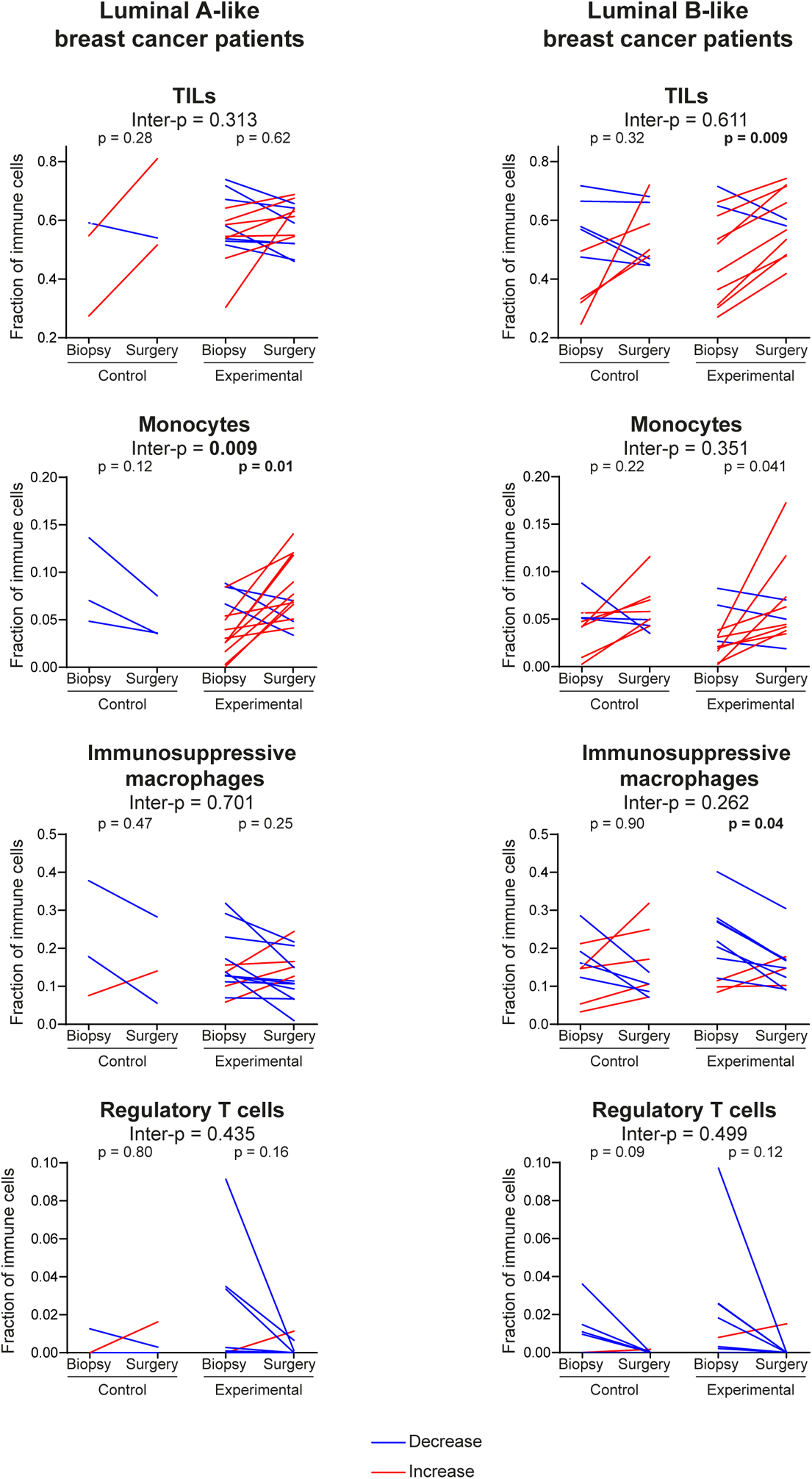
Denosumab immunomodulatory effects are more pronounced in luminal-B like tumours. CIBERSORT-inferred fractions of TILs, monocytes, immunosuppressive macrophages and regulatory T cells in paired biopsy and surgery samples from luminal A-like (control n = 3; denosumab n = 14) and luminal B-like (control n = 9; denosumab n = 11) patients with early-stage luminal breast cancer. Values are proportions from 0 to 1. Red and blue lines denote increases and decreases from biopsy to surgery, respectively. P values above paired plots are from paired t-tests within each arm; inter-P values compare surgery-biopsy changes between denosumab and control arms by unpaired t-test.

**Extended Data Figure 4.**
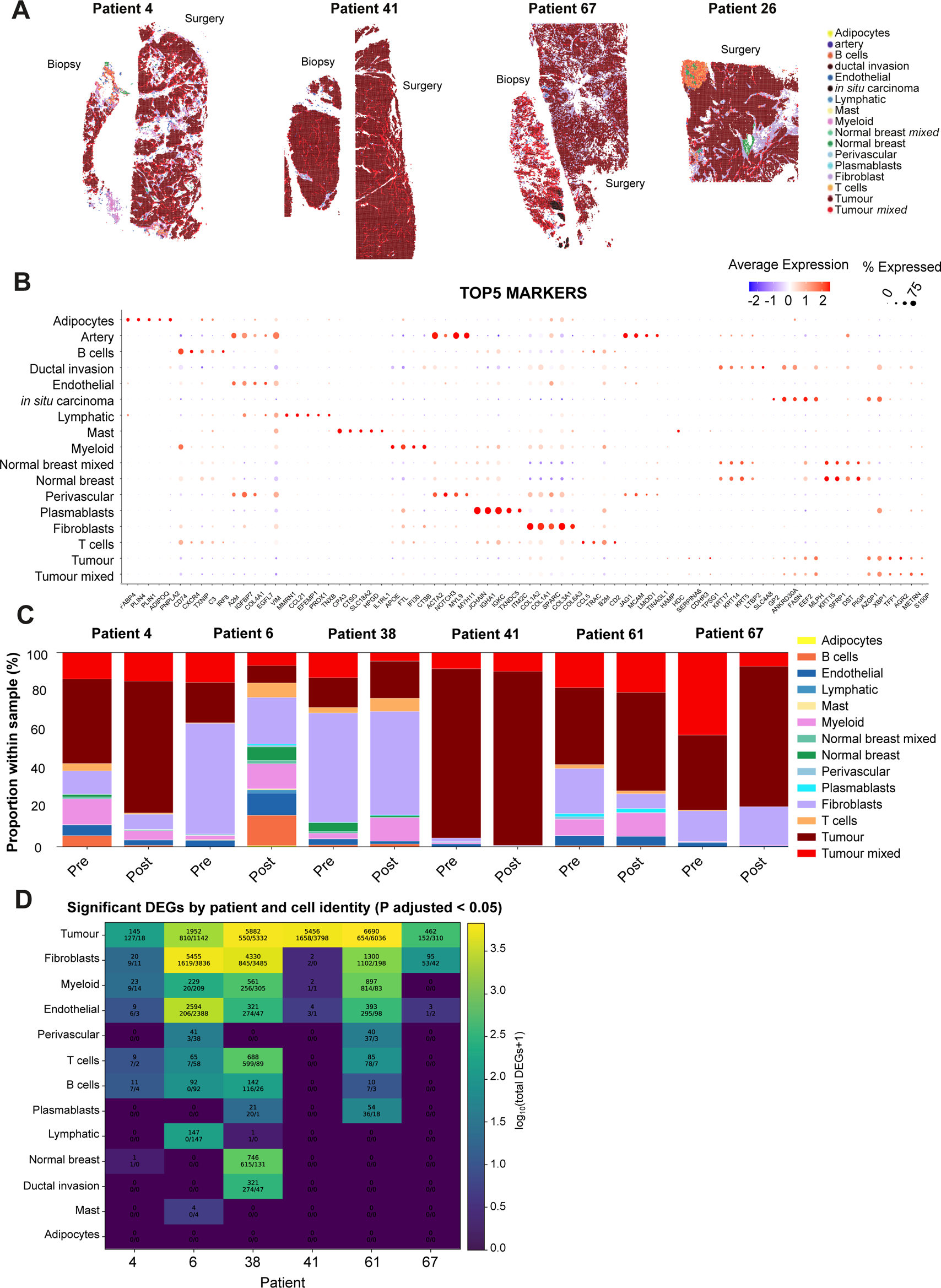
Validation of Visium HD annotation and patient-level spatial composition. A. Spatial maps of annotated Visium HD spots in biopsy and surgery sections from patients 4, 41 and 67 coloured by final cell identity. Only a surgery sample was available for patient 26. B. Dot plot showing the top five marker genes for each annotated Visium HD population. Dot colour indicates scaled average expression, and dot size indicates the percentage of spots expressing the marker. Patient 26 was included for annotation validation but excluded from paired downstream analyses because no matched biopsy Visium HD sample was available. C. Stacked bar plots showing the proportion of annotated cell identities within each biopsy and surgery sample. D. Heatmap showing the number of significant DEGs for each patient and annotated cell identity in the patient-level Visium HD analysis. Colour indicates log^10^(number of DEGs + 1); significant DEGs were defined by adjusted p < 0.05.

**Extended Data Figure 5.**
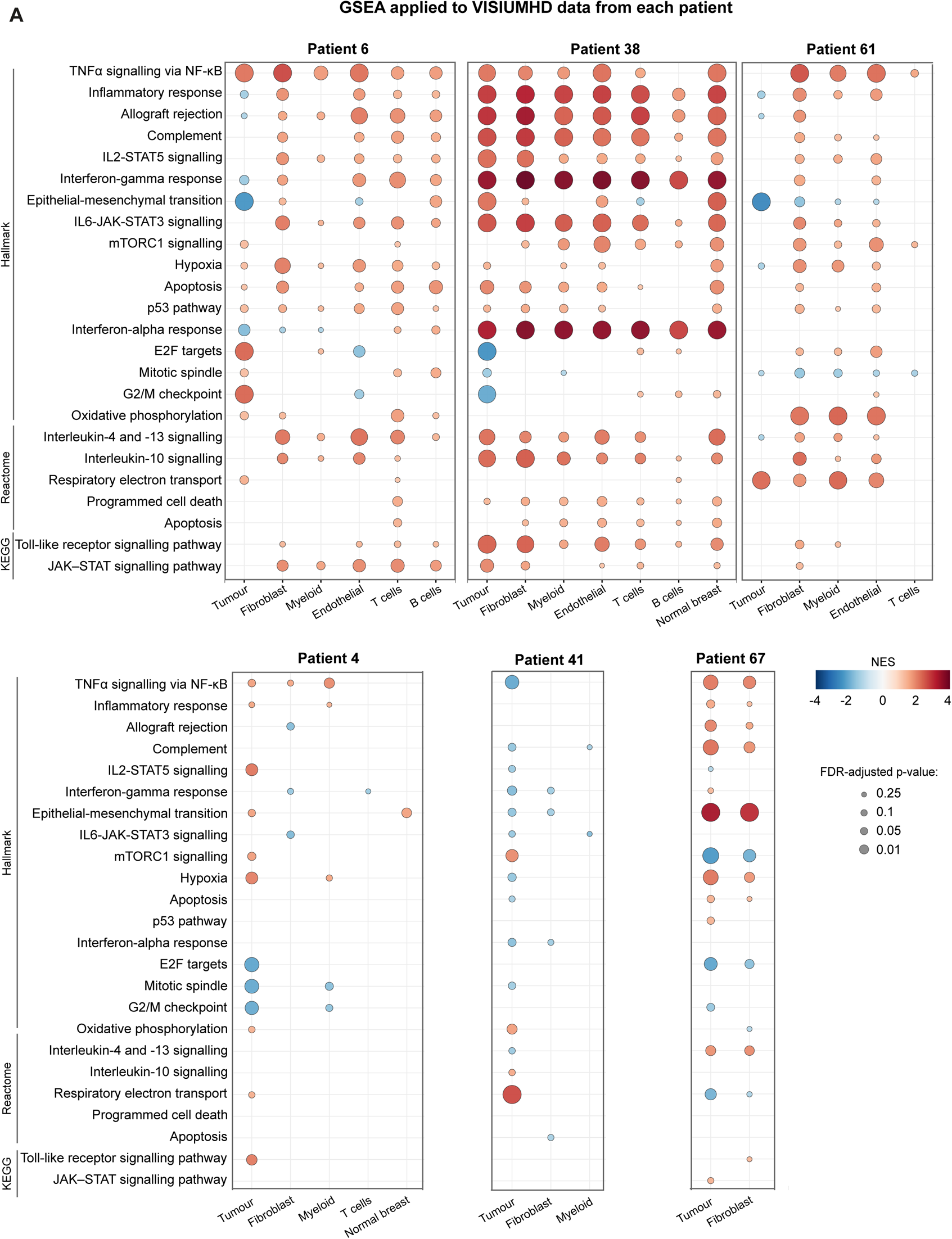
Patient-level GSEA reveals heterogeneous but recurrent transcriptional responses to denosumab. Bubble plot of GSEA results from patient-level Visium HD pseudobulk analyses across annotated cell compartments in patients 6, 38, 61, 4, 41 and 67. Dot colour indicates the NES and dot size indicates FDR-adjusted q-value. Positive NES values indicate surgery enrichment, and negative NES values indicate biopsy enrichment. Pathways are grouped by Hallmark, Reactome and KEGG collections.

**Extended Data Figure 6.**
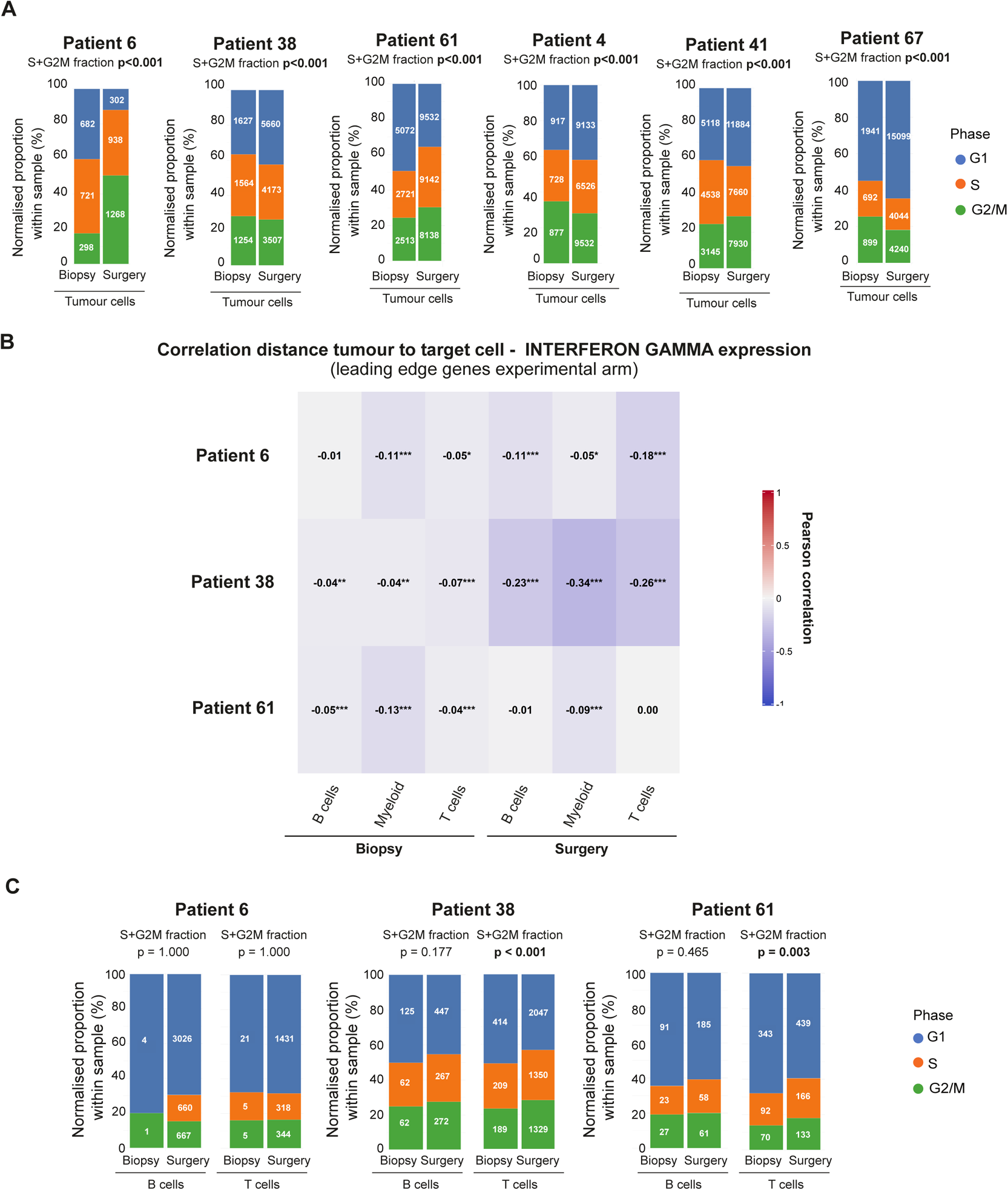
Cell-cycle state and interferon-distance relationships. A. Cell-cycle phase distribution in tumour cells from paired biopsy and surgery Visium HD samples across patients. Bars show normalised proportions of G1, S and G2/M phases within tumour cells, with cell counts shown within bars where applicable. Statistical annotations indicate S+G2M biopsy-surgery comparisons. Note that “cell division” decreases in tumour cells for patients 38, 4, 41 and 67. B. Relationship between the expression of selected denosumab-associated interferon-gamma signature genes (based on leading-edge genes) and distance from tumour regions in the indicated immune populations. Correlation coefficients (r) and p values are shown for biopsy and surgery samples. C. Cell-cycle phase distribution in B cell and T cell compartments from immune-informative Visium HD cases. Bars show normalised proportions of G1, S and G2/M phases in biopsy and surgery samples. P values for biopsy–surgery comparisons of the S + G2/M fraction are also shown.

**Extended Data Figure 7.**
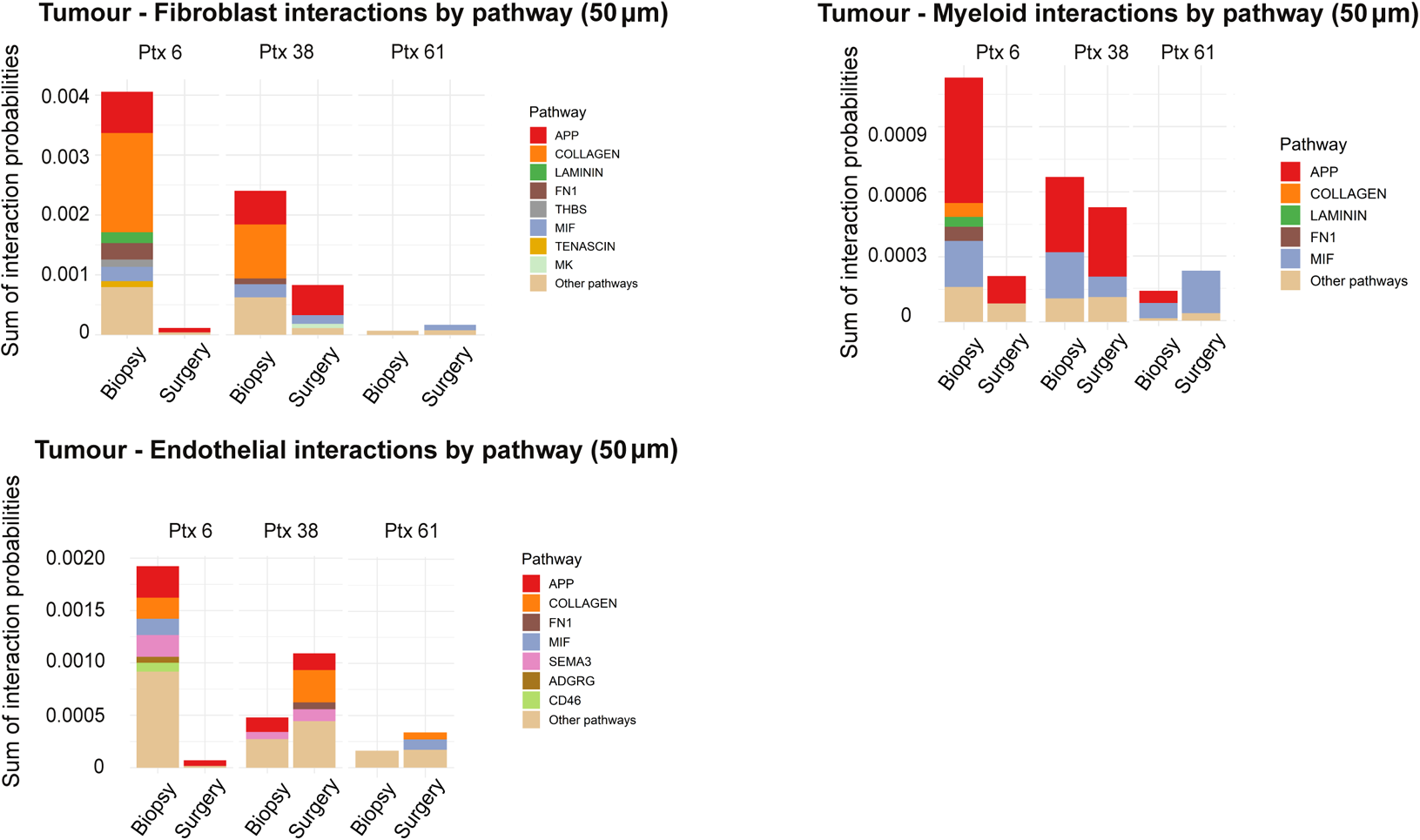
Pathway-level tumour-fibroblast, tumour-myeloid and tumour-endothelial communication after denosumab. CellChat-derived pathway-level interaction probabilities for tumour-fibroblast, tumour-myeloid and tumour-endothelial communication at a 50-μm neighbourhood interaction scale. Stacked bars show summed interaction probabilities for each signalling pathway before and after denosumab across the indicated patients. Colours indicate CellChat signalling categories.

